# Comparing the Diagnostic Performance of qPCR, ddPCR, and NGS Liquid Biopsies for HPV-Associated Cancers

**DOI:** 10.1101/2023.09.26.23296002

**Authors:** Saskia Naegele, Daniel A. Ruiz-Torres, Yan Zhao, Deborah Goss, Daniel L. Faden

## Abstract

HPV-associated cancers, including oropharyngeal squamous cell carcinoma(HPV+OPSCC), cervical cancer(HPV+CC), and squamous cell carcinoma of the anus(HPV+SCCA), release circulating tumor HPV DNA(ctHPVDNA) into the blood. The diagnostic performance of ctHPVDNA detection depends on the approaches utilized and the individual assay metrics. A comparison of these approaches has not been systematically performed to inform expected performance, which in turn impacts clinical interpretation. A meta-analysis was performed using Ovid MEDLINE, Embase, and Web of Science Core Collection databases to assess the diagnostic accuracy of ctHPVDNA detection across cancer anatomic sites, detection platforms, and blood components. The population included HPV+OPSCC, HPV+CC, and HPV+SCCA patients with pre-treatment samples analyzed by quantitative PCR(qPCR), digital droplet PCR(ddPCR), or next generation sequencing(NGS). 36 studies involving 2,986 patients met the inclusion criteria. The sensitivity, specificity and quality of each study were assessed and pooled for each analysis.

The sensitivity of ctHPVDNA detection was greatest with NGS, followed by ddPCR and lastly qPCR when pooling all studies, while specificity was similar(sensitivity: ddPCR>qPCR, p<0.001; NGS>ddPCR, p=0.014). ctHPVDNA from OPSCC was more easily detected compared to CC and SCCA, overall(p=0.044).

In conclusion, detection platform, anatomic site of the cancer and blood component utilized impacts ctHPVDNA detection and must be considered when interpreting results. Plasma NGS-based testing should be considered the most sensitive approach for ctHPVDNA overall.

## Introduction

Human papillomaviruses (HPVs) are a family of DNA oncoviruses that cause benign and malignant lesions of the genital mucosa, upper respiratory tract, and skin. More than 200 distinct types of HPV have been identified, and at least 14 of them are classified as high-risk, or capable of tumorigenesis, in specific anatomic sites^1,2,3^. HPV accounts for 5% of cancers worldwide and causes almost all cases of cervical cancer, as well as a significant proportion of vaginal, vulvar, penile, anal, rectal, and oropharyngeal cancers^4–8^. Routine screening strategies for HPV-associated cervical cancer (HPV+CC), including pelvic exams and Papanicolaou smears, enable early detection and have contributed to a substantial reduction in the incidence of early-stage cervical cancer^9,10^. Unlike cervical cancer, effective screening strategies for HPV-associated oropharyngeal squamous cell carcinoma (HPV+OPSCC) and HPV-associated squamous cell carcinoma of the anus (HPV+SCCA) are lacking, and the incidence of these cancers has steadily increased in the past few decades^11^. In the United States and the United Kingdom, the incidence of HPV+OPSCC in men has surpassed rates of cervical cancer in women and continues to rise, despite HPV vaccination efforts^12–16^. Existing approaches to diagnose and monitor these cancers are invasive, costly and have variable accuracy, indicating a need to improve the current standard of care.

Circulating tumor DNA (ctDNA) is released or secreted from cancer cells into the blood and other body fluids^17–22^. Liquid biopsies detecting ctDNA have demonstrated broad applicability from cancer screening to molecular profiling, adaptive treatment monitoring during therapy, detection of minimal residual disease, and recurrence detection^23–33^. HPV-associated cancers release ctHPVDNA, which has distinct advantages over somatic ctDNA due to the smaller size of the viral genome and its specificity to cancer cells, making HPV-associated cancers an optimal target for the application of liquid biopsies^34^. Numerous studies have demonstrated that ctHPVDNA is detectable in the plasma or serum of patients with HPV-associated cancers at the time of diagnosis, can be used as a real-time biomarker to monitor treatment response, and detects recurrence earlier than standard of care imaging^35–45^. ctHPVDNA-based diagnostics may have improved accuracy, reduced cost, and a shorter time to diagnosis compared with existing tissue-based clinical diagnostics^46^. In the past, conventional quantitative-polymerase chain reaction (qPCR) was the most common method employed for the detection of ctDNA, but newer (and more costly) techniques, including droplet-digital PCR (ddPCR) and next generation sequencing (NGS) have emerged^47^. The diagnostic performance of ctHPVDNA detection depends on the approaches utilized and the individual assay metrics. A comparison of these approaches has not been systematically performed in the literature to inform expected performance, which in turn impacts clinical interpretation. We sought to test the hypothesis that the sensitivity of NGS-based liquid biopsy is superior to qPCR and ddPCR at the time of diagnosis across HPV-associated cancers.

## Materials and Methods

A systematic review was performed by a medical librarian (D.G.) following the guidelines of the Preferred Reporting Items for Systematic Reviews and Meta-Analyses (PRISMA)^48^.

### Literature Search

A search of published studies in Ovid MEDLINE (1946-), Embase.com (1947-) and Web of Science Core Collections (1900-) was designed and conducted by a reference librarian (D.G.) on February 18 and 23, 2022. Search strategies were customized for each database (Supplementary Methods). Each search utilized a combination of controlled vocabulary and keyword terms relating to the diagnosis of HPV-associated cancer (head and neck, anal, vulvar, cervical, vaginal, penile) using qPCR, ddPCR, or NGS testing. The search was constructed to exclude non-human studies. No filters for language, study design, date of publication, or country of origin were used in the search producing 251 articles (Figure 1). All references were exported into EndNote x7.8. Duplicates were removed first by the automated process in EndNote and then manually by the librarian leaving 153 articles, which were exported into Covidence for study screening, selection, and data extraction. Nine subsequent articles were found through searching the references of included articles making up a total of 162 articles for screening.

**Figure 1.**
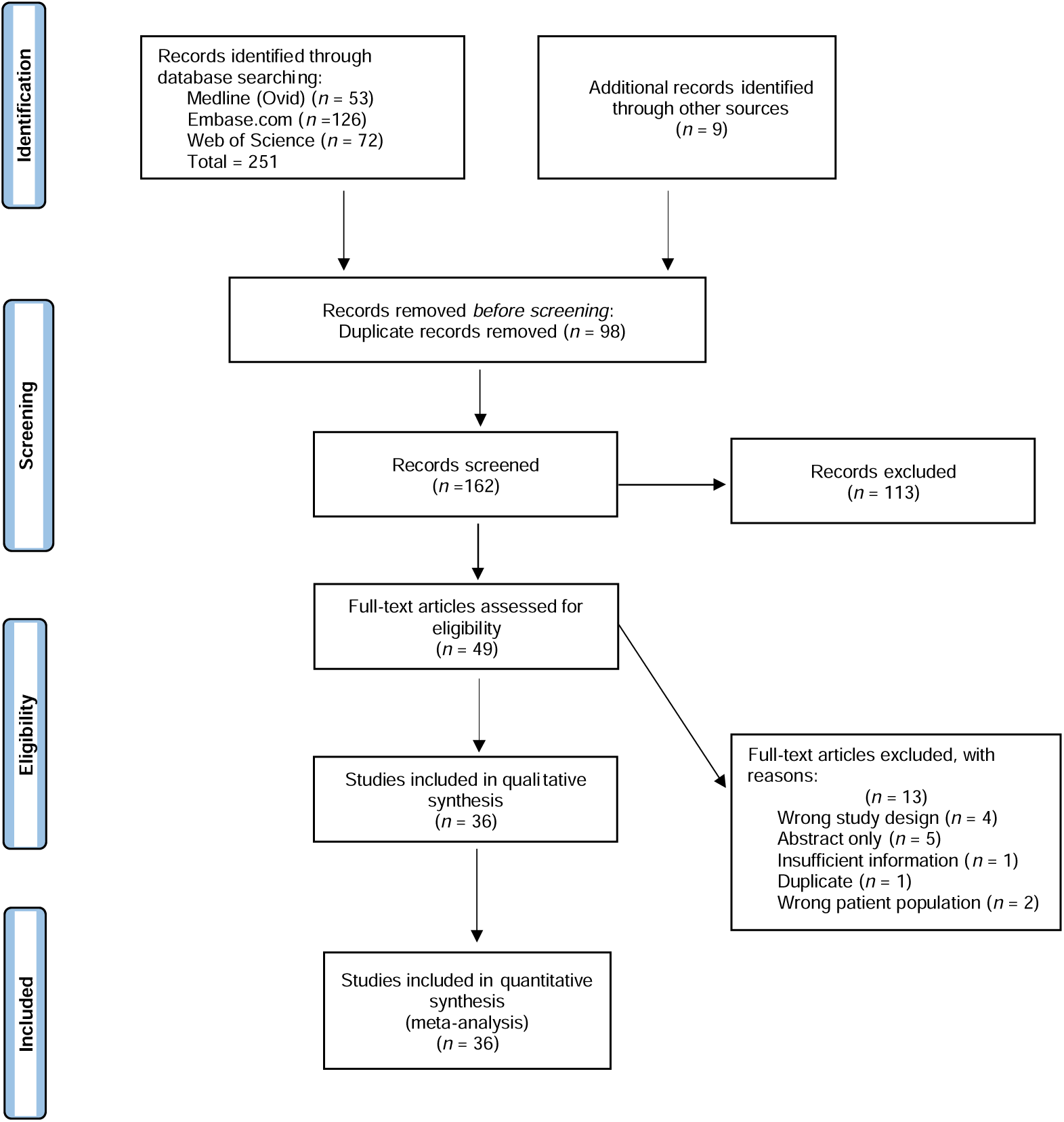
Preferred Reporting Items for Systematic Reviews and Meta-Analyses (PRISMA) flowchart depicting the study selection process.

### Study Selection

Studies that examined ctDNA in any HPV-associated cancer with qPCR, ddPCR, or NGS were considered eligible for inclusion. Extracted data comprised anatomic subsite, HPV status, HPV assay detection method, number of patients tested on each assay, number of true positives, false positives, false negatives, and true negatives, source of ctHPVDNA (plasma or serum), probe target gene (if ddPCR was used), and amplicon or hybrid capture (if NGS was used). Only studies written in English were included. Titles and abstracts were screened independently by two authors (S.N. and D.R.T.) for full text review. The same two authors independently conducted the full text review. Any disagreements in the screening process were settled by discussion and consensus between the two authors. All eligible studies were screened for duplicate data by comparing authors, timeframe of data collection, and outcomes. After full text screening, 36 studies remained for the quantitative synthesis.

R 4.1.2 was used to conduct the statistical analysis and the R packages “meta” and “metafor” were used for the meta-analysis. The studies missing one of the two values (TP, FN) or (TN, FP) were excluded. Using the random-effects model, sensitivity including 95% confidence intervals (CI) was computed from TP and TP + FN, and specificity including 95% CI was computed from TN and TN + FP. Subgroups were defined differently for each model and the pooled means were calculated respectively. Subsequently, separate meta-regressions were performed to test the association of each study characteristic with HPV sensitivities and specificities. Interstudy variability and between-study variance were assessed by Cochran’s Q statistic. The percentage of variation explained by true heterogeneity opposed to sampling error was calculated with I^2 statistic. We assume a two-sided p < 0.05 to be significant. Potential publication bias was evaluated using The Quality Assessment of Diagnostic Accuracy Studies second edition (QUADAS-2). The risk of bias was judged as high or low when the answers to all signaling questions in the four domains were yes or no, respectively. If the information was not sufficient, an unclear bias was used. Most studies were at unclear or low risk of bias for flow and timing and index test domains (Figure 2). Notably, for reference standard, three studies were at unclear risk of bias and for patient selection, four studies were at high risk of bias. Regarding applicability, 33 studies were at low risk of bias for reference standard and index test, but five studies were at high risk of bias for patient selection.

**Figure 2.**
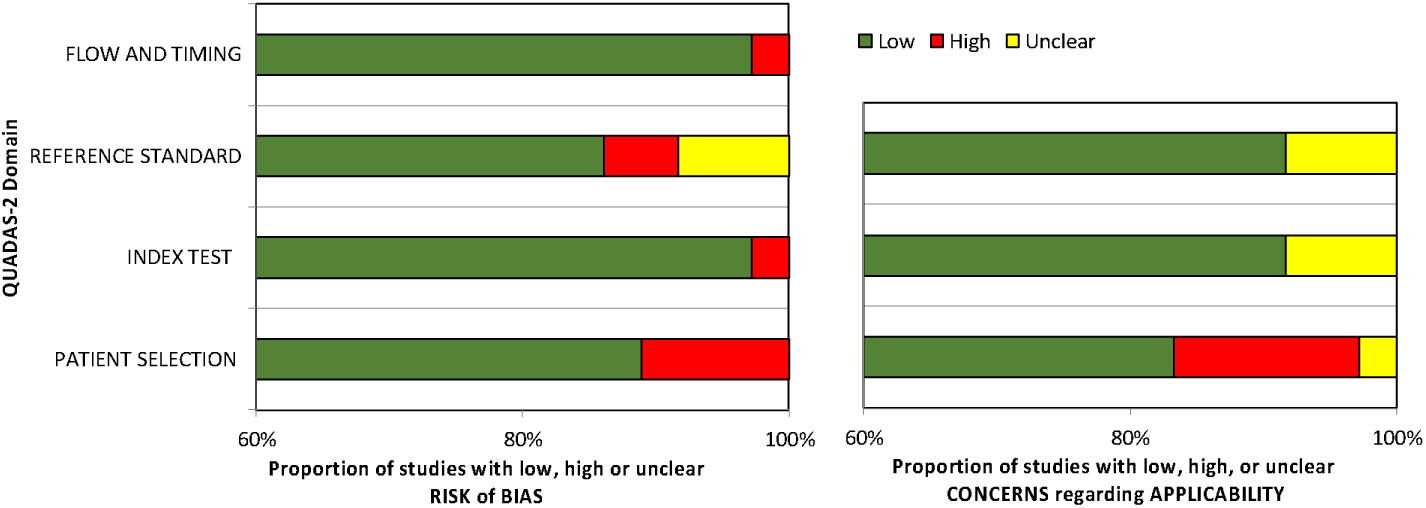
Quality evaluation of the included studies. Domains rated were flow and timing, patient selection, reference standard, and index test. Patient selection determined whether the selection of patients may have introduced bias. Index test assessed what the index test was and how it was conducted and interpreted. Reference standard assessed whether the gold standard was likely to correctly classify the target condition and if those results were interpreted without knowledge of the index test results. Flow and timing assessed whether there was an appropriate interval between the index test and the reference standard. Red color indicates high risk of bias, green indicates low risk of bias, and yellow indicates unclear risk of bias.

## Results

### Sensitivity and specificity of ctHPVDNA detection by test at the time of diagnosis

A total of 36 studies were included in the meta-analysis containing a total of 2,986 patients (Table 1 and Table S1). There were 11 qPCR studies, 19 ddPCR studies, and seven NGS studies. We first examined the pooled sensitivity of ctHPVDNA detection across all tests (Figure 3). We compared a pooled sensitivity of 0.81 (95% CI: 0.73-0.87) from 19 studies (n=1056) using ddPCR to 11 studies (n=597) using qPCR (0.51 (95% CI: 0.37-0.64), (p<0.001)) and seven studies (n=179) using NGS (0.94 (95% CI: 0.88-0.97), (p=0.014)) (Figure 4). 10 qPCR studies (n=638), 12 ddPCR studies (n=449), and seven NGS studies (n=244) were used to calculate specificity. A pooled specificity of 0.98 (95% CI: 0.96-0.99) for ddPCR was compared to qPCR (0.93 (95% CI: 0.83-0.97), (p=0.05)) and NGS (0.95 (95% CI: 0.90-0.97), (p=0.507)) (Figure 4).

**Figure 3.**
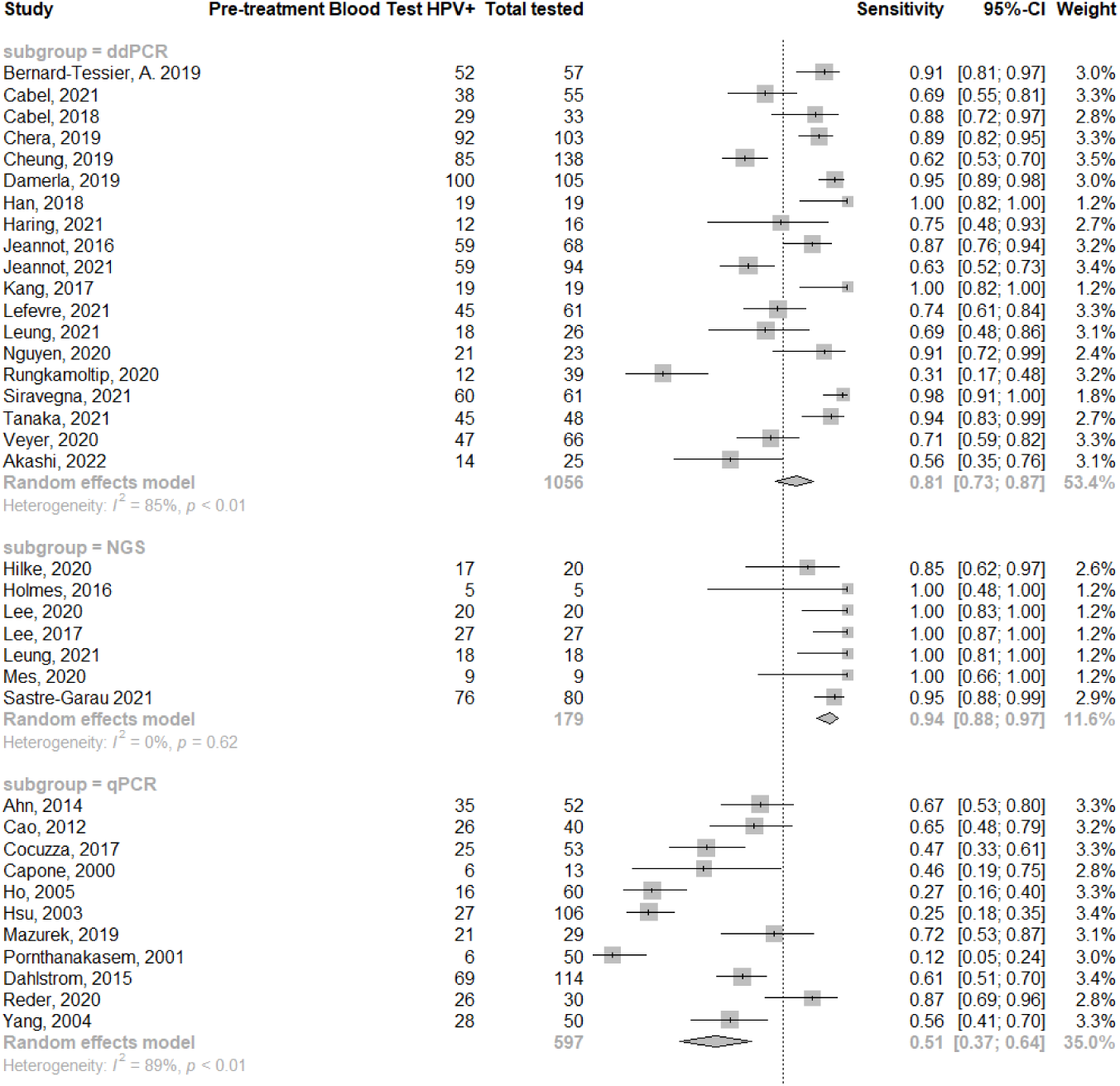
Sensitivity from 36 studies used in the meta-analysis.

**Figure 4.**
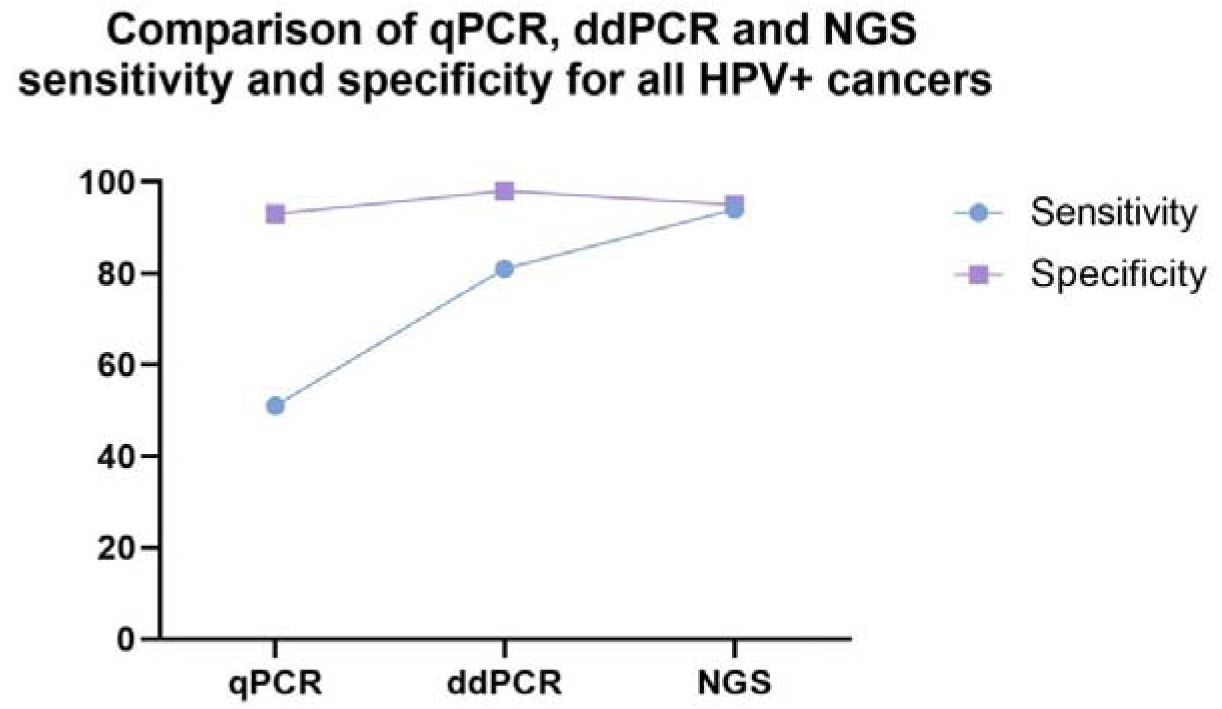
Pooled sensitivity and specificity of qPCR vs. ddPCR vs. NGS across all HPV-associated cancers.

**Table 1.**
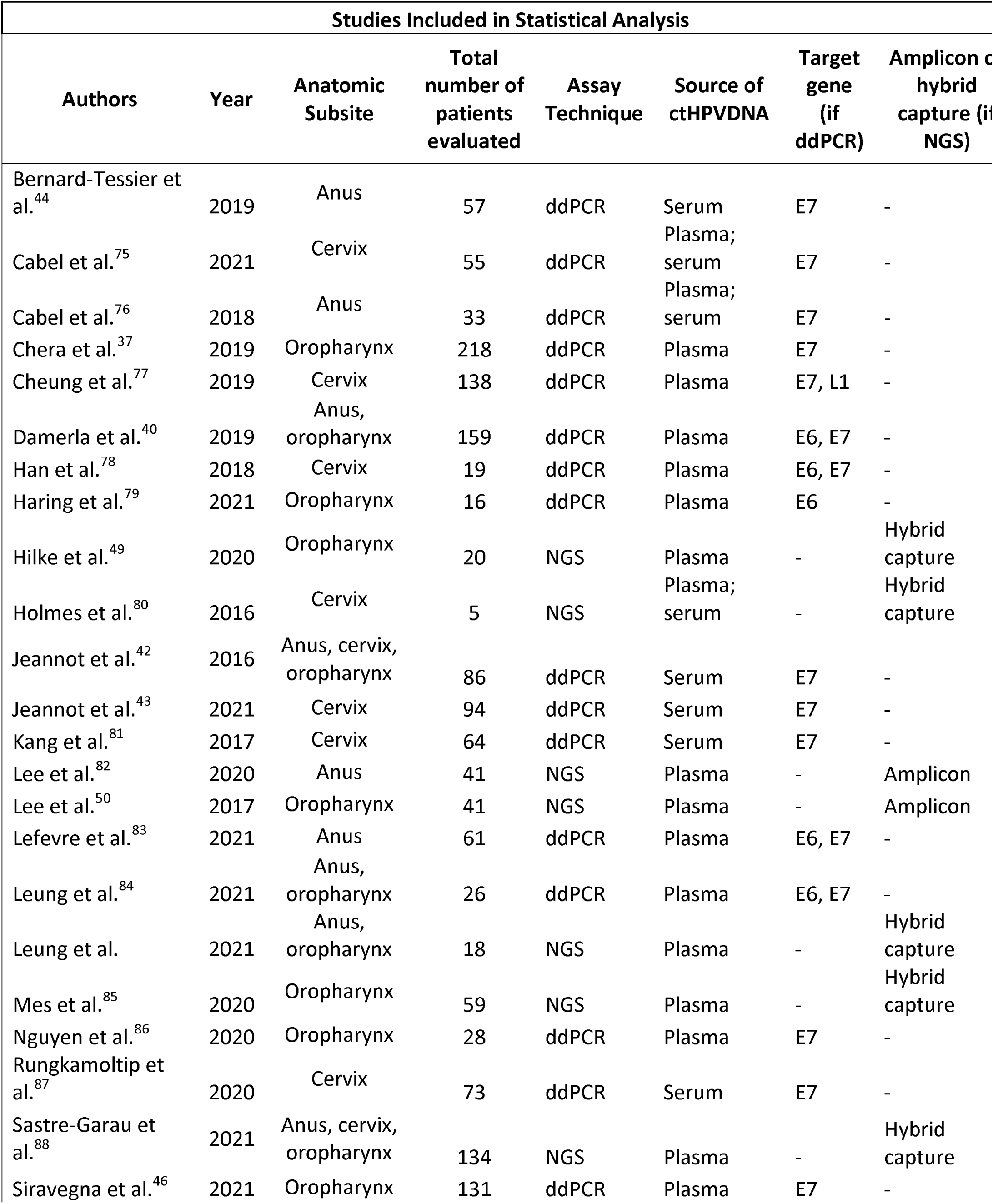
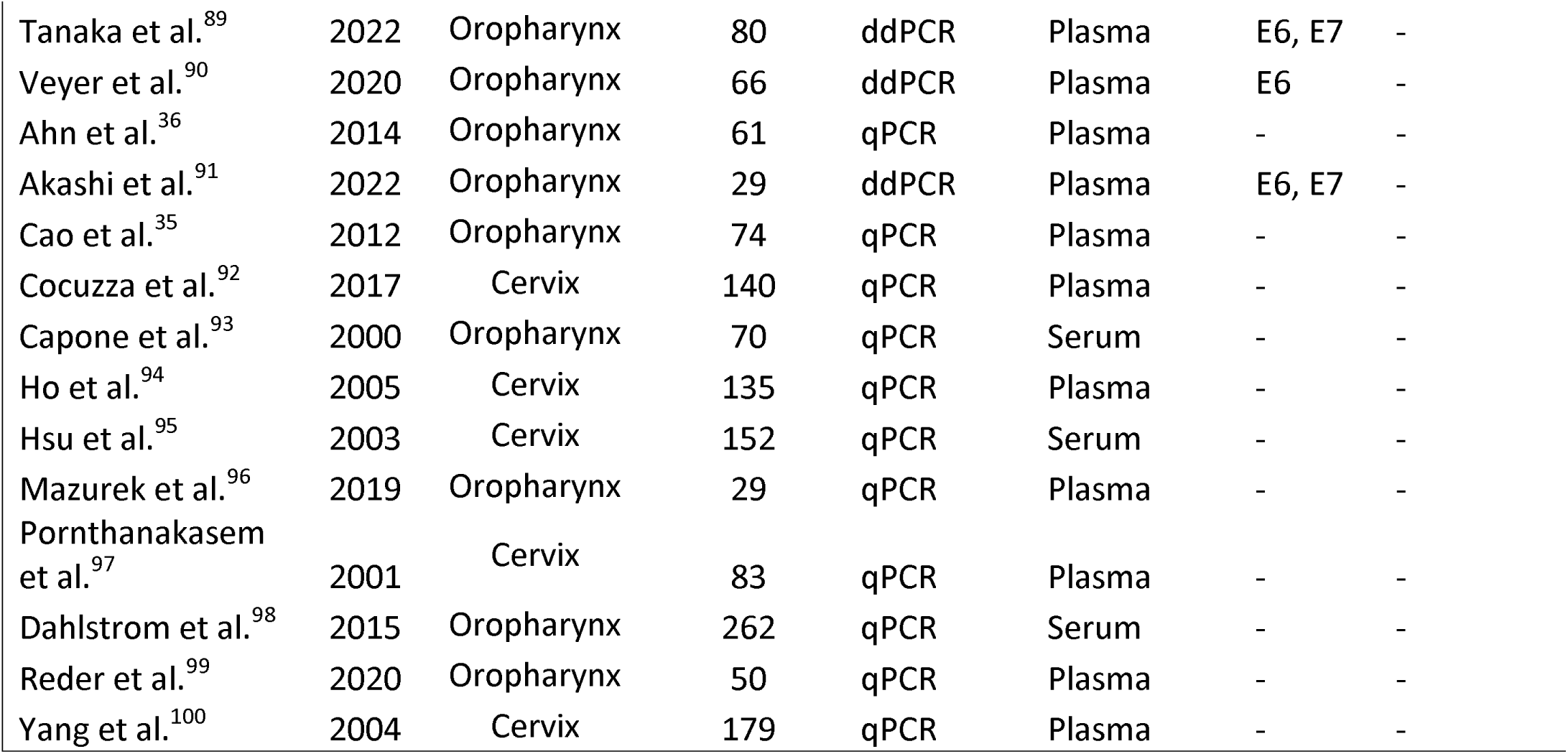
Overview of study characteristics. Table showing study characteristics for all 36 studies eligible for the statistical analysis including anatomic subsite, assay technique, source of ctHPVDNA, target gene (if ddPCR), and amplicon or hybrid capture (if NGS). “-“ indicates the characteristic was not applicable to the particular study.

### Sensitivity and specificity of ctHPVDNA detection test by anatomic subsite

As ctHPVDNA detection may differ by cancer anatomic site, we next evaluated sensitivity and specificity across each anatomic site by ctHPVDNA test. For HPV+OPSCC, we evaluated 21 studies containing a total of 1436 patients (Figure S1). We compared a pooled sensitivity of 0.89 (95% CI: 0.78-0.94) from 10 studies (n=460) using ddPCR to six studies (n=278) using qPCR (0.66 (95% CI: 0.58-0.74), (p=0.005)) and five studies (n=74) using NGS (0.91 (95% CI: 0.81-0.96), (p=0.357)) (Figure 5). Five qPCR studies (n=268), six ddPCR studies (n=253), and three NGS studies (n=103) were used to calculate specificity. A pooled specificity of 0.97 (95% CI: 0.94-0.99) for ddPCR was compared to qPCR (0.94 (95% CI: 0.59-0.99), (p=0.449)) and NGS (0.97 (95% CI: 0.90-0.99), (p=0.922)) (Figure 5).

**Figure 5.**
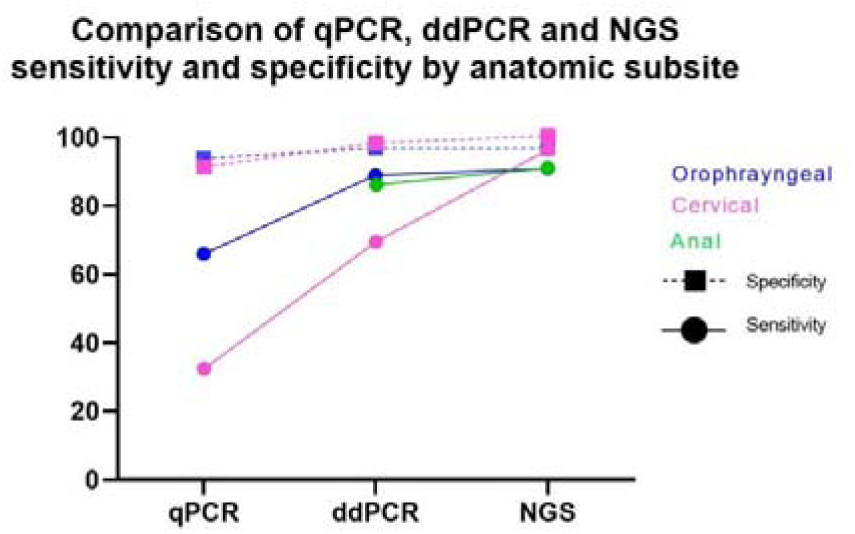
Pooled sensitivity and specificity of qPCR vs. ddPCR vs. NGS in studies of HPV-associated oropharyngeal, cervical, and anal cancer.

For HPV+CC we evaluated 16 studies containing a total of 1285 patients (Figure S2). We compared a pooled sensitivity of 0.69 (95% CI: 0.55-0.80) from eight studies (n=416) using ddPCR to five studies (n=319) using qPCR (0.32 (95% CI: 0.19-0.48), (p<0.001)) and three studies (n=72) using NGS (0.96 (95% CI: 0.87-0.99), (p=0.008)) (Figure 6). Five qPCR studies (n=370), three ddPCR studies (n=97), and one NGS study (n=11) were used to calculate specificity. A pooled specificity of 0.98 (95% CI: 0.92-1.00) for ddPCR was compared to qPCR (0.91 (95% CI: 0.80-0.96), (p=0.057)) and NGS (1.00 (95% CI: 0.72-1.00), (p=0.589)) (Figure 5).

**Figure 6.**
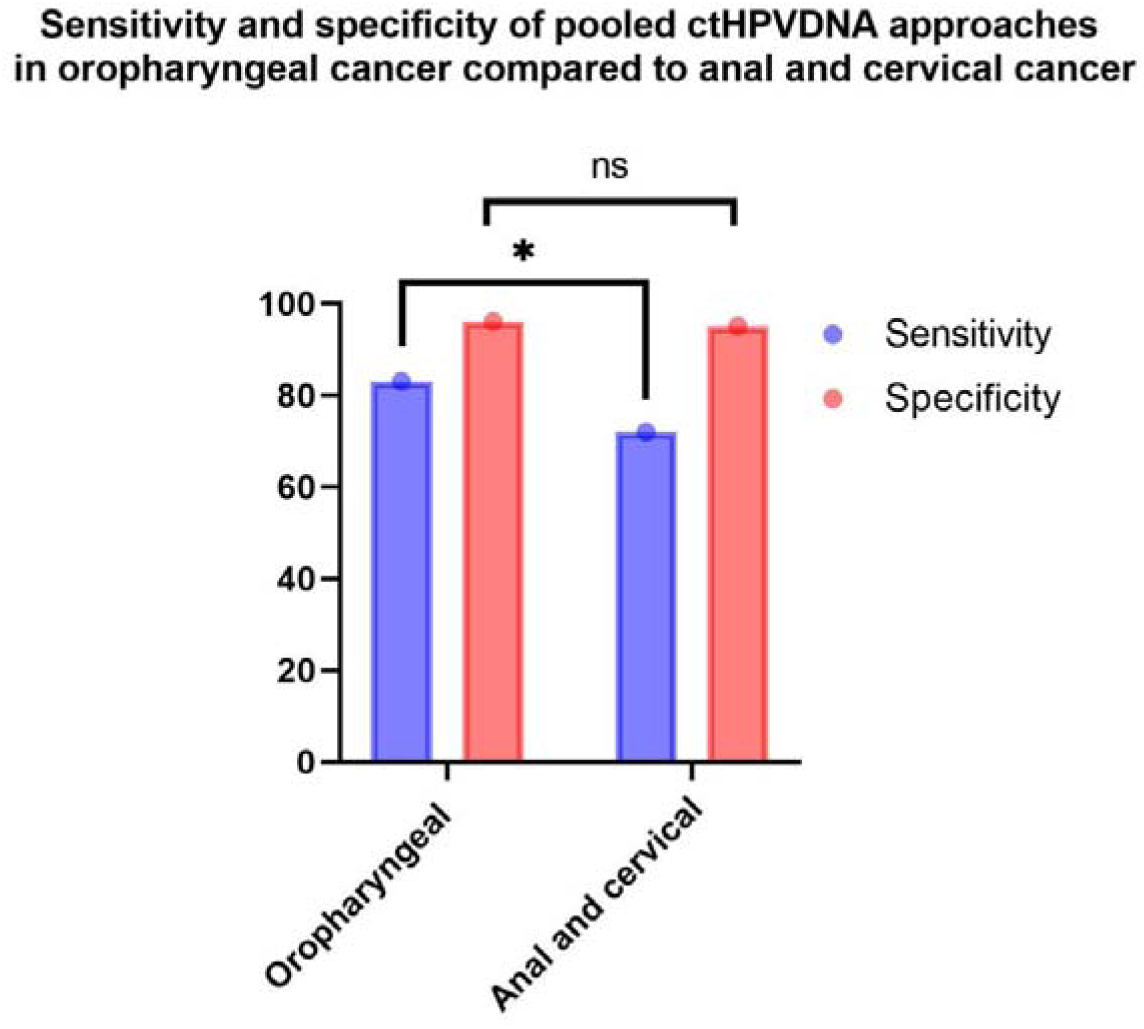
Pooled qPCR, ddPCR, and NGS sensitivity and specificity in studies of HPV-associated oropharyngeal cancer compared to cervical and anal cancer. “*” indicates a statistically significant p-value. “ns” indicates a nonsignificant p-value.

For HPV+SCCA we evaluated seven studies containing a total of 252 patients (Figure S3). We compared a pooled sensitivity of 0.86 (95% CI: 0.75-0.92) from five studies (n=172) using ddPCR to 0.91 (95% CI: 0.59-0.99) from two studies (n=32) using NGS (p=0.652) (Figure 5). Specificity regression was unavailable for HPV+SCCA studies because of the small sample size.

Lastly, we compared the pooled sensitivities and specificities of these assays in the most common HPV-associated cancer, HPV+OPSCC, which has distinct histopathologic features, to HPV-associated cervical and anal cancer (Figure S4, S5, S6, S7). We compared a pooled sensitivity of 0.66 (95% CI: 0.58-0.74) from six HPV+OPSCC qPCR studies (n= 278) to 0.32 (95% CI: 0.19-0.48) from five HPV-associated cervical and anal cancer qPCR studies (n=319) (p<0.001); a pooled sensitivity of 0.89 (95% CI: 0.78-0.94) from 10 HPV+OPSCC ddPCR studies (n=460) was compared to 0.77 (95% CI: 0.67-0.85) from 13 HPV-associated cervical and anal cancer ddPCR studies (n=588) (p=0.096); and a pooled sensitivity of 0.91 (95% CI: 0.81-0.96) from five HPV+OPSCC NGS studies (n=74) was compared to 0.93 (95% CI: 0.86-0.97) from five HPV-associated cervical and anal cancer studies (n=104) (p=0.645). Finally, a pooled sensitivity of 0.83 (95% CI: 0.75-0.89) from 21 HPV+OPSCC studies (n=812) using qPCR, ddPCR, and NGS was compared to a pooled sensitivity of 0.72 (95% CI: 0.60-0.81) from 23 HPV-associated anal and cervical cancer studies (n=1,011) using all three assays (p=0.044) (Figure 6).

A pooled specificity of 0.94 (95% CI: 0.59-0.99) from five HPV+OPSCC qPCR studies (n=268) was compared to 0.91 (95% CI: 0.80-0.96) from five HPV-associated cervical and anal cancer qPCR studies (n=370) (p=0.710). A pooled specificity of 0.97 (95% CI: 0.94-0.99) from six HPV+OPSCC ddPCR studies (n=253) was compared to 0.98 (95% CI: 0.94-1.00) from four HPV-associated cervical and anal cancer ddPCR studies (n=124) (p=0.489). A pooled specificity of 0.97 (95% CI: 0.90-0.99) from three HPV+OPSCC NGS studies (n=103) was compared to 0.97 (95% CI: 0.81-1.00) from two HPV-associated cervical and anal cancer NGS studies (n=32). Finally, a pooled specificity of 0.96 (95% CI: 0.86-0.99) from 14 HPV+OPSCC studies (n=624) using qPCR, ddPCR, and NGS was compared to a pooled specificity of 0.95 (95% CI: 0.90-0.98) from 11 HPV-associated anal and cervical cancer studies (n=526) using all three assays (p=0.738) (Figure 6).

### Sensitivity and specificity of test by blood compartment

We next evaluated the pooled sensitivities and specificities based on whether the studies used plasma or serum samples (Figure S8, S9, S10, S11) to evaluate the impact of the blood compartment on test performance. A pooled sensitivity of 0.65 (95% CI: 0.45-0.81) from eight serum qPCR, ddPCR, and NGS studies combined (n=510) was compared to 0.79 (95% CI: 0.70-0.86) from 27 plasma studies (n=1240) using all three assays (p=0.125). A pooled specificity of 0.97 (95% CI: 0.70-1.00) from six serum qPCR, ddPCR, and NGS studies combined (n=348) was compared to 0.95 (95% CI: 0.91-0.97) from 18 plasma studies (n=861) using all three assays (p=0.840).

### Sensitivity and specificity of test by target

We evaluated the pooled sensitivities and specificities based on the targets of the three assays (Figure S12). First, we looked at ddPCR studies which had probes designed for a single E7 target gene and compared them to ddPCR studies with probes for both the E6 and E7 genes. We compared a pooled sensitivity of 0.81 (95% CI: 0.69-0.89) from 11 studies (n=690) using E7 ddPCR probes to 0.84 (95% CI: 0.67-0.93) from six studies (n=284) which used both E6 and E7 ddPCR probes (p=0.727). A pooled specificity of 0.98 (95% CI: 0.95-0.99) from six studies (n=287) whose ddPCR assays targeted E7 was compared to 0.97 (95% CI: 0.89-0.99) from three studies (n=90) which used both E6 and E7 ddPCR probes (p=0.732).

Next, we evaluated NGS studies based on whether they used an amplicon-based approach or hybrid capture (Figure S13). A pooled sensitivity of 0.93 (95% CI: 0.86-0.96) from five NGS hybrid capture studies (n=132) was compared to 0.98 (95% CI: 0.87-1.00) from two NGS amplicon studies (n=47) (p=0.224). A pooled specificity of 0.96 (95% CI: 0.83-0.99) from three NGS hybrid capture studies (n=159) was then compared to 0.95 (95% CI: 0.79-0.99) from two NGS amplicon studies (n=35) (p=0.955). Of the seven NGS studies, five of these used assays that covered the full genome. One of the studies, Hilke et al., only covered E7 and another study, Lee et al., only covered 40% of the genome, focusing on sub-lineage defining regions^49,50^. It is notable that in the two amplicon-based studies, whole genome versus partial genome coverage did not have a difference in sensitivity whereas in the hybrid capture studies, the one study that only covered the E7 region had a noticeably lower sensitivity (0.85) compared to the overall pooled sensitivity (0.93).

## Discussion

Despite advances in early detection for HPV+CC, including Pap smears and direct HPV PCR testing, cervical cancer remains the fourth-leading cause of cancer death in women worldwide and is the leading cause of cancer death in women in low-human development index countries^51–53^. For HPV+SCCA, there has been a dramatic increase in the incidence and mortality rates (nearly 3% per year) with roughly 10,000 new cases and over 1,600 deaths expected to be attributed to it in in the United States in 2022^11,54–56^. Globally, high– and middle-income countries have detected a 2% to 6% annual increase in the HPV+SCCA incidence over the last few decades^57,58^. Similarly concerning, and even more striking, the incidence of HPV+OPSCC has risen more than 200% over the past decades in the United States and is projected to continue climbing, despite HPV vaccination efforts^12,15,16,59,60^.

Blood-based biomarkers function as ideal, non-invasive modalities with strong diagnostic and monitoring potential for HPV-associated cancers. Accurate detection of ctHPVDNA has the potential to optimize many aspects of cancer management, from the pre-diagnostic setting to minimal residual disease (MRD) detection after surgery and detection of recurrence. As HPV-associated cancers remain a global health concern, and as the field of ctHPVDNA detection continues to evolve, it is critical to have a better understanding of the available ctHPVDNA detection approaches and how they perform both against each other and amongst differing anatomic subsites. In this meta-analysis, we examined 36 studies to compare the sensitivities and specificities across qPCR, ddPCR, and NGS platforms in HPV+OPSCC, SCCA, and CC at the time of diagnosis.

After pooling the sensitivities from 36 studies, we found that NGS was the most sensitive platform across all cancer types, outperforming ddPCR, which similarly outperformed qPCR.

Why NGS is the most sensitive approach for ctHPVDNA detection is likely multifaceted. First, current ddPCR and qPCR approaches are only able to detect a limited number of pre-defined targets. Considering the HPV genome is ∼8,000 base pairs, targeting one or two ∼160 base pair fragments (the approximate size of cell free DNA) leaves about 99% of the genome untargeted. Thus, while NGS capture is less efficient than ddPCR, the significant increase in the number of targets overcomes this limitation while leading to improved sensitivity. Further, while the majority of HPV+OPSCC, SCCA, and CC are caused by a limited number of high-risk HPV genotypes, ∼5% of cases are caused by numerous other rare genotypes, which are missed by ddPCR and qPCR approaches, further limiting sensitivity^61–64^. Lastly, ddPCR and qPCR approaches are subject to false negatives due to mutations in the single DNA fragment of interest, disrupting primer binding, which is a phenomenon we have seen in our lab in select cases.

Similarly, ddPCR was more sensitive than qPCR. The reason for ddPCR’s superiority over qPCR lies in its inherent technological design. ddPCR partitions each sample into thousands of individual oil-water emulsion droplets, each of which are then individually analyzed for the presence or absence of a fluorescent signal. A crucial element of ddPCR is its ability to quantify the absolute number of DNA copies in each sample, without relying on a standard curve^65^. At low copy numbers, ddPCR affords greater sensitivity over qPCR because high-copy templates and background are diluted across the droplets, enhancing template concentration in HPV-positive partitions.

When comparing ctHPVDNA detection between anatomic sites, we discovered that across all platforms, detection was improved in the oropharynx compared to the anus and cervix. We believe there are a few reasons for the superiority of liquid biopsy in the oropharynx. First, and most critically, HPV specifically targets the palatine and lingual tonsils in the oropharynx, which contain branching tonsillar crypts. These crypts are lined by a highly specialized lymphoepithelium, known as the reticulated epithelium, which functions to transport foreign antigens from the surrounding environment of the oropharynx to the tonsillar lymphoid tissue. The porous nature of the basal cell layer of this epithelium permits the direct passage of lymphocytes and antigen-presenting cells. While the disrupted reticular epithelium plays a large role in mucosal immune protection, these same gaps also enable direct exposure to viral particles like HPV. Whereas in the cervix, HPV infection requires microtrauma, such as mechanical abrasion, of the epithelium with subsequent invasion of the virus through an exposed basement membrane, in the reticulated epithelium of the tonsil, the already porous epithelium exposes the basal cell layer and basement membrane to viral transgression without the need for mucosal disruption^60,66–69^. We hypothesize that just as the gaps in the basement membrane of the tonsil permit HPV infection and subsequent malignant transformation, they may provide a similar route for egress of ctHPVDNA from HPV+OPSCC, providing liquid biopsy assays with more ctDNA to detect compared to ctDNA from the cervix and anus. Second, HPV+OPSCC is often detected with nodal metastases^60^. Part of the reason for the early nodal metastasis of HPV+OPSCC may also be a result of the microanatomy of the crypt epithelium, which enables the cancer to easily spread to regional lymph nodes. The discontinuous basement membrane permits both early transgression of the cancer as well as regional metastasis of occult cancers. The nodal disease seen at presentation in most HPV+OPSCC cases means there is more tumor burden at diagnosis, which in turn leads to more shedding of ctHPVDNA, enhancing the sensitivity of HPV+OPSCC liquid biopsy at presentation. Importantly, existing data has shown that lymph node burden correlates most strongly with ctDNA levels. In a study of 110 patients, Rettig et al. found that the few patients with undetectable ctHPVDNA predominantly had clinical stage N0 disease^70^. This suggests that assays used at the time of diagnosis may have lower sensitivity among patients without regional lymph node metastasis. Since most HPV+OPSCC patients initially present with cervical lymphadenopathy due to asymptomatic early disease, liquid biopsy assays will thus have increased sensitivity for these patients.

The fact that ctHPVDNA may be more easily detected in HPV+OPSCC than CC or SCCA is further borne out when analyzing each HPV-associated cancer individually. For HPV+OPSCC, we discovered that while ddPCR was more sensitive than qPCR, the difference between ddPCR and NGS was not statistically significant, indicating that marginal improvements in sensitivity from ddPCR to NGS may not be meaningful when ctHPVDNA is relatively plentiful. On the contrary, when examining HPV+CC, where ctHPVDNA levels may be quantitatively lower at presentation due to both earlier cancer stage at presentation and less ctHPVDNA transgression into the circulation, NGS was more sensitive than ddPCR^71–74^. The power of a more sensitive diagnostic test is more important in HPV+CC because of this, whereas in HPV+OPSCC, the most sensitive technique is less critical, rendering the difference in sensitivity between NGS and ddPCR negligible.

This study has a number of limitations which relate to the status of the existing literature as well as the inherent methodological variation within the included studies. First, sample sizes of available studies are small, with 13 of the 36 studies included in the analysis having a sample size of under 50 patients. Second, there is significant heterogeneity of the molecular characteristics of each of these liquid biopsy assays, such as probe design, cell free DNA (cfDNA) input, and the thresholds for positive detection. Specifically for NGS, assays also differ markedly in their library design, preparation steps, depth of sequencing, and variation in downstream sequencing analysis. More broadly, variation of the cfDNA extraction process, volume of plasma extracted from, extraction kits and procedure, and the volume and concentration of cfDNA added to the assay all impact assay performance. Controlling for all of these variables in a meta-analysis would not be possible. Reassuringly, variation in cfDNA extraction is likely negligible compared to other factors, both biological and technical. An additional limitation of this meta-analysis is that the data output (HPV reads) is not normalized between studies and how this value is calculated is variable from assay to assay. We also did not evaluate the sensitivity and specificities across the different assays and anatomic sites by stage, which could affect our hypothesis that liquid biopsies targeting ctHPVDNA from the oropharynx have higher sensitivity due to late-stage disease presentation with increased N-stage and associated tumor burden. Lastly, because of the more recent emergence of NGS, there were fewer NGS studies for evaluation.

## Conclusion

In summary, using a systematic review and meta-analysis, detection platform, anatomic site of the cancer and blood component utilized for cell free DNA extraction impact ctHPVDNA detection and must be considered when interpreting ctHPVDNA test results. Currently, plasma NGS-based testing should be considered the most sensitive approach for ctHPVDNA detection overall, while specificity is excellent regardless of platform utilized.

## Data Availability

All data produced in the present study are available upon reasonable request to the authors.

## References

1. Saraiya M, Unger ER, Thompson TD, Lynch CF, Hernandez BY, Lyu CW, Steinau M, Watson M, Wilkinson EJ, Hopenhayn C, Copeland G, Cozen W, Peters ES, Huang Y, Saber MS, Altekruse S, Goodman MT. US assessment of HPV types in cancers: implications for current and 9-valent HPV vaccines. J Natl Cancer Inst. 2015;107(6):djv086.

2. Bzhalava D, Guan P, Franceschi S, Dillner J, Clifford G. A systematic review of the prevalence of mucosal and cutaneous human papillomavirus types. Virology. 2013;445(1-2):224–231.

3. Bouvard V, Baan R, Straif K, Grosse Y, Secretan B, El Ghissassi F, Benbrahim-Tallaa L, Guha N, Freeman C, Galichet L, Cogliano V. A review of human carcinogens--Part B: biological agents. Lancet Oncol. 2009;10(4):321–322.

4. de Martel C, Plummer M, Vignat J, Franceschi S. Worldwide burden of cancer attributable to HPV by site, country and HPV type. Int J Cancer. 2017;141(4):664–670.

5. Goodman MT, Saraiya M, Thompson TD, Steinau M, Hernandez BY, Lynch CF, Lyu CW, Wilkinson EJ, Tucker T, Copeland G, Peters ES, Altekruse S, Unger ER. Human papillomavirus genotype and oropharynx cancer survival in the United States of America. Eur J Cancer. 2015;51(18):2759–2767.

6. Alemany L, Saunier M, Alvarado-Cabrero I, Quirós B, Salmeron J, Shin HR, Pirog EC, Guimerà N, Hernandez-Suarez G, Felix A, Clavero O, Lloveras B, Kasamatsu E, Goodman MT, Hernandez BY, Laco J, Tinoco L, Geraets DT, Lynch CF, Mandys V, Poljak M, Jach R, Verge J, Clavel C, Ndiaye C, Klaustermeier J, Cubilla A, Castellsagué X, Bravo IG, Pawlita M, Quint WG, Muñoz N, Bosch FX, de Sanjosé S. Human papillomavirus DNA prevalence and type distribution in anal carcinomas worldwide. Int J Cancer. 2015;136(1):98–107.

7. Forman D, de Martel C, Lacey CJ, Soerjomataram I, Lortet-Tieulent J, Bruni L, Vignat J, Ferlay J, Bray F, Plummer M, Franceschi S. Global burden of human papillomavirus and related diseases. Vaccine. 2012;30 Suppl 5:F12–23.

8. Baricevic I, He X, Chakrabarty B, Oliver AW, Bailey C, Summers J, Hampson L, Hampson I, Gilbert DC, Renehan AG. High-sensitivity human papilloma virus genotyping reveals near universal positivity in anal squamous cell carcinoma: different implications for vaccine prevention and prognosis. Eur J Cancer. 2015;51(6):776–785.

9. Peirson L, Fitzpatrick-Lewis D, Ciliska D, Warren R. Screening for cervical cancer: a systematic review and meta-analysis. Syst Rev. 2013;2:35.

10. Whitlock EP, Vesco KK, Eder M, Lin JS, Senger CA, Burda BU. Liquid-based cytology and human papillomavirus testing to screen for cervical cancer: a systematic review for the U.S. Preventive Services Task Force. Ann Intern Med. 2011;155(10):687–697, w214-685.

11. Deshmukh AA, Suk R, Shiels MS, Sonawane K, Nyitray AG, Liu Y, Gaisa MM, Palefsky JM, Sigel K. Recent Trends in Squamous Cell Carcinoma of the Anus Incidence and Mortality in the United States, 2001-2015. J Natl Cancer Inst. 2020;112(8):829–838.

12. Lechner M, Jones OS, Breeze CE, Gilson R. Gender-neutral HPV vaccination in the UK, rising male oropharyngeal cancer rates, and lack of HPV awareness. Lancet Infect Dis. 2019;19(2):131–132.

13. Division of Cancer Prevention and Control CfDCaP: Number of HPV-Associated Cancer Cases per Year, CDC, 2019.

14. Tota JE, Best AF, Zumsteg ZS, Gillison ML, Rosenberg PS, Chaturvedi AK. Evolution of the Oropharynx Cancer Epidemic in the United States: Moderation of Increasing Incidence in Younger Individuals and Shift in the Burden to Older Individuals. Journal of clinical oncology: official journal of the American Society of Clinical Oncology. 2019;37(18):1538–1546.

15. Zhang Y, Fakhry C, D’Souza G. Projected Association of Human Papillomavirus Vaccination With Oropharynx Cancer Incidence in the US, 2020-2045. JAMA Oncol. 2021;7(10):e212907.

16. Chaturvedi AK, Engels EA, Pfeiffer RM, Hernandez BY, Xiao W, Kim E, Jiang B, Goodman MT, Sibug-Saber M, Cozen W, Liu L, Lynch CF, Wentzensen N, Jordan RC, Altekruse S, Anderson WF, Rosenberg PS, Gillison ML. Human papillomavirus and rising oropharyngeal cancer incidence in the United States. Journal of clinical oncology: official journal of the American Society of Clinical Oncology. 2011;29(32):4294–4301.

17. Siravegna G, Marsoni S, Siena S, Bardelli A. Integrating liquid biopsies into the management of cancer. Nat Rev Clin Oncol. 2017;14(9):531–548.

18. Cescon DW, Bratman SV, Chan SM, Siu LL. Circulating tumor DNA and liquid biopsy in oncology. Nat Cancer. 2020;1(3):276–290.

19. Wan JCM, Massie C, Garcia-Corbacho J, Mouliere F, Brenton JD, Caldas C, Pacey S, Baird R, Rosenfeld N. Liquid biopsies come of age: towards implementation of circulating tumour DNA. Nat Rev Cancer. 2017;17(4):223–238.

20. Crowley E, Di Nicolantonio F, Loupakis F, Bardelli A. Liquid biopsy: monitoring cancer-genetics in the blood. Nat Rev Clin Oncol. 2013;10(8):472–484.

21. Pascual J, Attard G, Bidard FC, Curigliano G, De Mattos-Arruda L, Diehn M, Italiano A, Lindberg J, Merker JD, Montagut C, Normanno N, Pantel K, Pentheroudakis G, Popat S, Reis-Filho JS, Tie J, Seoane J, Tarazona N, Yoshino T, Turner NC. ESMO recommendations on the use of circulating tumour DNA assays for patients with cancer: a report from the ESMO Precision Medicine Working Group. Annals of oncology: official journal of the European Society for Medical Oncology. 2022;33(8):750–768.

22. Thierry AR, El Messaoudi S, Gahan PB, Anker P, Stroun M. Origins, structures, and functions of circulating DNA in oncology. Cancer Metastasis Rev. 2016;35(3):347–376.

23. Dawson SJ, Tsui DW, Murtaza M, Biggs H, Rueda OM, Chin SF, Dunning MJ, Gale D, Forshew T, Mahler-Araujo B, Rajan S, Humphray S, Becq J, Halsall D, Wallis M, Bentley D, Caldas C, Rosenfeld N. Analysis of circulating tumor DNA to monitor metastatic breast cancer. N Engl J Med. 2013;368(13):1199–1209.

24. Chen K, Zhao H, Shi Y, Yang F, Wang LT, Kang G, Nie Y, Wang J. Perioperative Dynamic Changes in Circulating Tumor DNA in Patients with Lung Cancer (DYNAMIC). Clinical cancer research: an official journal of the American Association for Cancer Research. 2019;25(23):7058–7067.

25. Bettegowda C, Sausen M, Leary RJ, Kinde I, Wang Y, Agrawal N, et al. Detection of circulating tumor DNA in early#x2013; and late-stage human malignancies. Sci Transl Med. 2014;6(224):224ra224.

26. Stejskal P, Goodarzi H, Srovnal J, Hajdúch M, van’t Veer LJ, Magbanua MJM. Circulating tumor nucleic acids: biology, release mechanisms, and clinical relevance. Molecular Cancer. 2023;22(1):15.

27. Bernard V, Kim DU, San Lucas FA, Castillo J, Allenson K, Mulu FC, Stephens BM, Huang J, Semaan A, Guerrero PA, Kamyabi N, Zhao J, Hurd MW, Koay EJ, Taniguchi CM, Herman JM, Javle M, Wolff R, Katz M, Varadhachary G, Maitra A, Alvarez HA. Circulating Nucleic Acids Are Associated With Outcomes of Patients With Pancreatic Cancer. Gastroenterology. 2019;156(1):108–118.e104.

28. Li RY, Liang ZY. Circulating tumor DNA in lung cancer: real-time monitoring of disease evolution and treatment response. Chin Med J (Engl). 2020;133(20):2476–2485.

29. Magbanua MJM, Swigart LB, Wu HT, Hirst GL, Yau C, Wolf DM, Tin A, Salari R, Shchegrova S, Pawar H, Delson AL, DeMichele A, Liu MC, Chien AJ, Tripathy D, Asare S, Lin CJ, Billings P, Aleshin A, Sethi H, Louie M, Zimmermann B, Esserman LJ, van’t Veer LJ. Circulating tumor DNA in neoadjuvant-treated breast cancer reflects response and survival. Ann Oncol. 2021;32(2):229–239.

30. Schwarzenbach H, Alix-Panabières C, Müller I, Letang N, Vendrell JP, Rebillard X, Pantel K. Cell-free tumor DNA in blood plasma as a marker for circulating tumor cells in prostate cancer. Clin Cancer Res. 2009;15(3):1032–1038.

31. Tie J, Wang Y, Tomasetti C, Li L, Springer S, Kinde I, Silliman N, Tacey M, Wong HL, Christie M, Kosmider S, Skinner I, Wong R, Steel M, Tran B, Desai J, Jones I, Haydon A, Hayes T, Price TJ, Strausberg RL, Diaz LA, Jr., Papadopoulos N, Kinzler KW, Vogelstein B, Gibbs P. Circulating tumor DNA analysis detects minimal residual disease and predicts recurrence in patients with stage II colon cancer. Sci Transl Med. 2016;8(346):346ra392.

32. Tie J, Cohen JD, Wang Y, Christie M, Simons K, Lee M, Wong R, Kosmider S, Ananda S, McKendrick J, Lee B, Cho JH, Faragher I, Jones IT, Ptak J, Schaeffer MJ, Silliman N, Dobbyn L, Li L, Tomasetti C, Papadopoulos N, Kinzler KW, Vogelstein B, Gibbs P. Circulating Tumor DNA Analyses as Markers of Recurrence Risk and Benefit of Adjuvant Therapy for Stage III Colon Cancer. JAMA Oncol. 2019;5(12):1710–1717.

33. Reinert T, Henriksen TV, Christensen E, Sharma S, Salari R, Sethi H, et al. Analysis of Plasma Cell-Free DNA by Ultradeep Sequencing in Patients With Stages I to III Colorectal Cancer. JAMA Oncol. 2019;5(8):1124–1131.

34. Faden DL. Liquid biopsy for the diagnosis of HPV-associated head and neck cancer. Cancer cytopathology. 2022;130(1):12–15.

35. Cao H, Banh A, Kwok S, Shi X, Wu S, Krakow T, Khong B, Bavan B, Bala R, Pinsky BA, Colevas D, Pourmand N, Koong AC, Kong CS, Le QT. Quantitation of human papillomavirus DNA in plasma of oropharyngeal carcinoma patients. International journal of radiation oncology, biology, physics. 2012;82(3):e351–358.

36. Ahn SM, Chan JY, Zhang Z, Wang H, Khan Z, Bishop JA, Westra W, Koch WM, Califano JA. Saliva and plasma quantitative polymerase chain reaction-based detection and surveillance of human papillomavirus-related head and neck cancer. JAMA Otolaryngol Head Neck Surg. 2014;140(9):846–854.

37. Chera BS, Kumar S, Beaty BT, Marron D, Jefferys S, Green R, Goldman EC, Amdur R, Sheets N, Dagan R, Hayes DN, Weiss J, Grilley-Olson JE, Zanation A, Hackman T, Blumberg JM, Patel S, Weissler M, Tan XM, Parker JS, Mendenhall W, Gupta GP. Rapid Clearance Profile of Plasma Circulating Tumor HPV Type 16 DNA during Chemoradiotherapy Correlates with Disease Control in HPV-Associated Oropharyngeal Cancer. Clin Cancer Res. 2019;25(15):4682–4690.

38. Chera BS, Kumar S, Shen C, Amdur R, Dagan R, Green R, Goldman E, Weiss J, Grilley-Olson J, Patel S, Zanation A, Hackman T, Blumberg J, Patel S, Thorp B, Weissler M, Yarbrough W, Sheets N, Mendenhall W, Tan XM, Gupta GP. Plasma Circulating Tumor HPV DNA for the Surveillance of Cancer Recurrence in HPV-Associated Oropharyngeal Cancer. Journal of clinical oncology: official journal of the American Society of Clinical Oncology. 2020;38(10):1050–1058.

39. Hanna GJ, Lau CJ, Mahmood U, Supplee JG, Mogili AR, Haddad RI, Janne PA, Paweletz CP. Salivary HPV DNA informs locoregional disease status in advanced HPV-associated oropharyngeal cancer. Oral oncology. 2019;95:120–126.

40. Damerla RR, Lee NY, You D, Soni R, Shah R, Reyngold M, Katabi N, Wu V, McBride SM, Tsai CJ, Riaz N, Powell SN, Babady NE, Viale A, Higginson DS. Detection of Early Human Papillomavirus-Associated Cancers by Liquid Biopsy. JCO Precis Oncol. 2019;3.

41. Naegele S, Efthymiou V, Das D, Sadow PM, Richmon JD, Iafrate AJ, Faden DL. Detection and Monitoring of Circulating Tumor HPV DNA in HPV-Associated Sinonasal and Nasopharyngeal Cancers. JAMA Otolaryngol Head Neck Surg. 2022.

42. Jeannot E, Becette V, Campitelli M, Calméjane MA, Lappartient E, Ruff E, Saada S, Holmes A, Bellet D, Sastre-Garau X. Circulating human papillomavirus DNA detected using droplet digital PCR in the serum of patients diagnosed with early stage human papillomavirus-associated invasive carcinoma. J Pathol Clin Res. 2016;2(4):201–209.

43. Jeannot E, Latouche A, Bonneau C, Calméjane MA, Beaufort C, Ruigrok-Ritstier K, Bataillon G, Larbi Chérif L, Dupain C, Lecerf C, Popovic M, de la Rochefordière A, Lecuru F, Fourchotte V, Jordanova ES, von der Leyen H, Tran-Perennou C, Legrier ME, Dureau S, Raizonville L, Bello Roufai D, Le Tourneau C, Bièche I, Rouzier R, Berns E, Kamal M, Scholl S. Circulating HPV DNA as a Marker for Early Detection of Relapse in Patients with Cervical Cancer. Clin Cancer Res. 2021;27(21):5869–5877.

44. Bernard-Tessier A, Jeannot E, Guenat D, Debernardi A, Michel M, Proudhon C, Vincent-Salomon A, Bièche I, Pierga JY, Buecher B, Meurisse A, François É, Cohen R, Jary M, Vendrely V, Samalin E, El Hajbi F, Baba-Hamed N, Borg C, Bidard FC, Kim S. Clinical Validity of HPV Circulating Tumor DNA in Advanced Anal Carcinoma: An Ancillary Study to the Epitopes-HPV02 Trial. Clin Cancer Res. 2019;25(7):2109–2115.

45. Naegele S, Efthymiou V, Hirayama S, Zhao BY, Das D, Chan AW, Richmon JD, Iafrate AJ, Faden DL. Double trouble: Synchronous and metachronous primaries confound ctHPVDNA monitoring. Head Neck. 2023.

46. Siravegna G, O’Boyle CJ, Varmeh S, Queenan N, Michel A, Stein J, Thierauf J, Sadow PM, Faquin WC, Perry SK, Bard AZ, Wang W, Deschler DG, Emerick KS, Varvares MA, Park JC, Clark JR, Chan AW, Andreu Arasa VC, Sakai O, Lennerz J, Corcoran RB, Wirth LJ, Lin DT, Iafrate AJ, Richmon JD, Faden DL. Cell free HPV DNA provides an accurate and rapid diagnosis of HPV-associated head and neck cancer. Clinical Cancer Research. 2021:clincanres.3151.2021.

47. Postel M, Roosen A, Laurent-Puig P, Taly V, Wang-Renault SF. Droplet-based digital PCR and next generation sequencing for monitoring circulating tumor DNA: a cancer diagnostic perspective. Expert Rev Mol Diagn. 2018;18(1):7–17.

48. Moher D, Liberati A, Tetzlaff J, Altman DG. Preferred reporting items for systematic reviews and meta-analyses: the PRISMA statement. Ann Intern Med. 2009;151(4):264–269, w264.

49. Hilke FJ, Muyas F, Admard J, Kootz B, Nann D, Welz S, Rieß O, Zips D, Ossowski S, Schroeder C, Clasen K. Dynamics of cell-free tumour DNA correlate with treatment response of head and neck cancer patients receiving radiochemotherapy. Radiother Oncol. 2020;151:182–189.

50. Lee JY, Garcia-Murillas I, Cutts RJ, De Castro DG, Grove L, Hurley T, Wang F, Nutting C, Newbold K, Harrington K, Turner N, Bhide S. Predicting response to radical (chemo)radiotherapy with circulating HPV DNA in locally advanced head and neck squamous carcinoma. Br J Cancer. 2017;117(6):876–883.

51. American Cancer Society. Global Cancer Facts & Figures 4th Edition. Atlanta: American Cancer Society; 2018.

52. Sung H, Ferlay J, Siegel RL, Laversanne M, Soerjomataram I, Jemal A, Bray F. Global Cancer Statistics 2020: GLOBOCAN Estimates of Incidence and Mortality Worldwide for 36 Cancers in 185 Countries. CA Cancer J Clin. 2021;71(3):209–249.

53. Wild CP, Weiderpass E, Stewart BW, editors (2020). World Cancer Report: Cancer Research for Cancer Prevention. Lyon, France: International Agency for Research on Cancer.

54. Damgacioglu H, Lin Y-Y, Ortiz AP, Wu C-F, Shahmoradi Z, Shyu SS, Li R, Nyitray AG, Sigel K, Clifford GM, Jay N, Lopez VC, Barnell GM, Chiao EY, Stier EA, Ortiz-Ortiz KJ, Ramos-Cartagena JM, Sonawane K, Deshmukh AA. State Variation in Squamous Cell Carcinoma of the Anus Incidence and Mortality, and Association With HIV/AIDS and Smoking in the United States. Journal of Clinical Oncology.0(0):JCO.22.01390.

55. Ortiz-Ortiz KJ, Ramos-Cartagena JM, Deshmukh AA, Torres-Cintrón CR, Colón-López V, Ortiz AP. Squamous Cell Carcinoma of the Anus Incidence, Mortality, and Survival Among the General Population and Persons Living With HIV in Puerto Rico, 2000-2016. JCO Glob Oncol. 2021;7:133–143.

56. Deshmukh AA, Suk R, Shiels MS, Damgacioglu H, Lin YY, Stier EA, Nyitray AG, Chiao EY, Nemutlu GS, Chhatwal J, Schmeler K, Sigel K, Sonawane K. Incidence Trends and Burden of Human Papillomavirus-Associated Cancers Among Women in the United States, 2001-2017. J Natl Cancer Inst. 2021;113(6):792–796.

57. Deshmukh AA, Damgacioglu H, Georges D, Sonawane K, Ferlay J, Bray F, Clifford GM. Global burden of HPV-attributable squamous cell carcinoma of the anus in 2020, according to sex and HIV status: A worldwide analysis. Int J Cancer. 2023;152(3):417–428.

58. Islami F, Ferlay J, Lortet-Tieulent J, Bray F, Jemal A. International trends in anal cancer incidence rates. Int J Epidemiol. 2017;46(3):924–938.

59. Faraji F, Rettig EM, Tsai HL, El Asmar M, Fung N, Eisele DW, Fakhry C. The prevalence of human papillomavirus in oropharyngeal cancer is increasing regardless of sex or race, and the influence of sex and race on survival is modified by human papillomavirus tumor status. Cancer. 2019;125(5):761–769.

60. Lechner M, Liu J, Masterson L, Fenton TR. HPV-associated oropharyngeal cancer: epidemiology, molecular biology and clinical management. Nat Rev Clin Oncol. 2022;19(5):306–327.

61. Lin C, Franceschi S, Clifford GM. Human papillomavirus types from infection to cancer in the anus, according to sex and HIV status: a systematic review and meta-analysis. Lancet Infect Dis. 2018;18(2):198–206.

62. So KA, Lee IH, Lee KH, Hong SR, Kim YJ, Seo HH, Kim TJ. Human papillomavirus genotype-specific risk in cervical carcinogenesis. J Gynecol Oncol. 2019;30(4):e52.

63. Castellsagué X, Alemany L, Quer M, Halec G, Quirós B, Tous S, et al. HPV Involvement in Head and Neck Cancers: Comprehensive Assessment of Biomarkers in 3680 Patients. J Natl Cancer Inst. 2016;108(6):djv403.

64. Gillison ML, Akagi K, Xiao W, Jiang B, Pickard RKL, Li J, Swanson BJ, Agrawal AD, Zucker M, Stache-Crain B, Emde AK, Geiger HM, Robine N, Coombes KR, Symer DE. Human papillomavirus and the landscape of secondary genetic alterations in oral cancers. Genome Res. 2019;29(1):1–17.

65. Hindson BJ, Ness KD, Masquelier DA, Belgrader P, Heredia NJ, Makarewicz AJ, et al. High-throughput droplet digital PCR system for absolute quantitation of DNA copy number. Anal Chem. 2011;83(22):8604–8610.

66. Roberts JN, Buck CB, Thompson CD, Kines R, Bernardo M, Choyke PL, Lowy DR, Schiller JT. Genital transmission of HPV in a mouse model is potentiated by nonoxynol-9 and inhibited by carrageenan. Nat Med. 2007;13(7):857–861.

67. Schiller JT, Day PM, Kines RC. Current understanding of the mechanism of HPV infection. Gynecol Oncol. 2010;118(1 Suppl):S12–17.

68. Ferris RL, Westra W. Oropharyngeal Carcinoma with a Special Focus on HPV-Related Squamous Cell Carcinoma. Annual Review of Pathology: Mechanisms of Disease. 2023;18(1):515–535.

69. Westra WH. The morphologic profile of HPV-related head and neck squamous carcinoma: implications for diagnosis, prognosis, and clinical management. Head Neck Pathol. 2012;6 Suppl 1(Suppl 1):S48–54.

70. Rettig EM, Wang AA, Tran NA, Carey E, Dey T, Schoenfeld JD, Sehgal K, Guenette JP, Margalit DN, Sethi R, Uppaluri R, Tishler RB, Annino DJ, Goguen LA, Jo VY, Haddad RI, Hanna GJ. Association of Pretreatment Circulating Tumor Tissue-Modified Viral HPV DNA With Clinicopathologic Factors in HPV-Positive Oropharyngeal Cancer. JAMA Otolaryngol Head Neck Surg. 2022;148(12):1120–1130.

71. Bui CN, Hong S, Suh M, Jun JK, Jung KW, Lim MC, Choi KS. Effect of Pap smear screening on cervical cancer stage at diagnosis: results from the Korean National Cancer Screening Program. J Gynecol Oncol. 2021;32(5):e81.

72. Landy R, Sasieni PD, Mathews C, Wiggins CL, Robertson M, McDonald YJ, Goldberg DW, Scarinci IC, Cuzick J, Wheeler CM. Impact of screening on cervical cancer incidence: A population-based case-control study in the United States. Int J Cancer. 2020;147(3):887–896.

73. Jun JK, Choi KS, Jung KW, Lee HY, Gapstur SM, Park EC, Yoo KY. Effectiveness of an organized cervical cancer screening program in Korea: results from a cohort study. Int J Cancer. 2009;124(1):188–193.

74. Landy R, Pesola F, Castañón A, Sasieni P. Impact of cervical screening on cervical cancer mortality: estimation using stage-specific results from a nested case-control study. Br J Cancer. 2016;115(9):1140–1146.

75. Cabel L, Bonneau C, Bernard-Tessier A, Héquet D, Tran-Perennou C, Bataillon G, Rouzier R, Féron JG, Fourchotte V, Le Brun JF, Benoît C, Rodrigues M, Scher N, Minsat M, Legrier ME, Bièche I, Proudhon C, Sastre-Garau X, Bidard FC, Jeannot E. HPV ctDNA detection of high-risk HPV types during chemoradiotherapy for locally advanced cervical cancer. ESMO Open. 2021;6(3):100154.

76. Cabel L, Jeannot E, Bieche I, Vacher S, Callens C, Bazire L, Morel A, Bernard-Tessier A, Chemlali W, Schnitzler A, Lièvre A, Otz J, Minsat M, Vincent-Salomon A, Pierga JY, Buecher B, Mariani P, Proudhon C, Bidard FC, Cacheux W. Prognostic Impact of Residual HPV ctDNA Detection after Chemoradiotherapy for Anal Squamous Cell Carcinoma. Clin Cancer Res. 2018;24(22):5767–5771.

77. Cheung TH, Yim SF, Yu MY, Worley MJ, Jr., Fiascone SJ, Chiu RWK, Lo KWK, Siu NSS, Wong MCS, Yeung ACM, Wong RRY, Chen ZG, Elias KM, Chung TKH, Berkowitz RS, Wong YF, Chan PKS. Liquid biopsy of HPV DNA in cervical cancer. J Clin Virol. 2019;114:32–36.

78. Han K, Leung E, Barbera L, Barnes E, Croke J, Grappa MAD, Fyles A, Metser U, Milosevic M, Pintilie M, Wolfson R, Zhao Z, Bratman SV. Circulating Human Papillomavirus DNA as a Biomarker of Response in Patients With Locally Advanced Cervical Cancer Treated With Definitive Chemoradiation. JCO Precision Oncology. 2018(2):1–8.

79. Haring CT, Bhambhani C, Brummel C, Jewell B, Bellile E, Heft Neal ME, Sandford E, Spengler RM, Bhangale A, Spector ME, McHugh J, Prince ME, Mierzwa M, Worden FP, Tewari M, Swiecicki PL, Brenner JC. Human papilloma virus circulating tumor DNA assay predicts treatment response in recurrent/metastatic head and neck squamous cell carcinoma. Oncotarget. 2021;12(13):1214–1229.

80. Holmes A, Lameiras S, Jeannot E, Marie Y, Castera L, Sastre-Garau X, Nicolas A. Mechanistic signatures of HPV insertions in cervical carcinomas. NPJ Genom Med. 2016;1:16004.

81. Kang Z, Stevanović S, Hinrichs CS, Cao L. Circulating Cell-free DNA for Metastatic Cervical Cancer Detection, Genotyping, and Monitoring. Clin Cancer Res. 2017;23(22):6856–6862.

82. Lee JY, Cutts RJ, White I, Augustin Y, Garcia-Murillas I, Fenwick K, Matthews N, Turner NC, Harrington K, Gilbert DC, Bhide S. Next Generation Sequencing Assay for Detection of Circulating HPV DNA (cHPV-DNA) in Patients Undergoing Radical (Chemo)Radiotherapy in Anal Squamous Cell Carcinoma (ASCC). Front Oncol. 2020;10:505.

83. Lefèvre AC, Pallisgaard N, Kronborg C, Wind KL, Krag SRP, Spindler KG. The Clinical Value of Measuring Circulating HPV DNA during Chemo-Radiotherapy in Squamous Cell Carcinoma of the Anus. Cancers (Basel). 2021;13(10).

84. Leung E, Han K, Zou J, Zhao Z, Zheng Y, Wang TT, Rostami A, Siu LL, Pugh TJ, Bratman SV. HPV Sequencing Facilitates Ultrasensitive Detection of HPV Circulating Tumor DNA. Clin Cancer Res. 2021.

85. Mes SW, Brink A, Sistermans EA, Straver R, Oudejans CBM, Poell JB, Leemans CR, Brakenhoff RH. Comprehensive multiparameter genetic analysis improves circulating tumor DNA detection in head and neck cancer patients. Oral Oncol. 2020;109:104852.

86. Nguyen B, Meehan K, Pereira MR, Mirzai B, Lim SH, Leslie C, Clark M, Sader C, Friedland P, Lindsay A, Tang C, Millward M, Gray ES, Lim AM. A comparative study of extracellular vesicle-associated and cell-free DNA and RNA for HPV detection in oropharyngeal squamous cell carcinoma. Scientific Reports. 2020;10(1):6083.

87. Rungkamoltip P, Temisak S, Piboonprai K, Japrung D, Thangsunan P, Chanpanitkitchot S, Chaowawanit W, Chandeying N, Tangjitgamol S, Iempridee T. Rapid and ultrasensitive detection of circulating human papillomavirus E7 cell-free DNA as a cervical cancer biomarker. Exp Biol Med (Maywood). 2021;246(6):654–666.

88. Sastre-Garau X, Diop M, Martin F, Dolivet G, Marchal F, Charra-Brunaud C, Peiffert D, Leufflen L, Dembélé B, Demange J, Tosti P, Thomas J, Leroux A, Merlin JL, Diop-Ndiaye H, Costa JM, Salleron J, Harlé A. A NGS-based Blood Test For the Diagnosis of Invasive HPV-associated Carcinomas with Extensive Viral Genomic Characterization. Clin Cancer Res. 2021;27(19):5307–5316.

89. Tanaka H, Suzuki M, Takemoto N, Fukusumi T, Eguchi H, Takai E, Kanai H, Tatsumi M, Horie M, Takenaka Y, Yachida S, Inohara H. Performance of oral HPV DNA, oral HPV mRNA and circulating tumor HPV DNA in the detection of HPV-related oropharyngeal cancer and cancer of unknown primary. Int J Cancer. 2022;150(1):174–186.

90. Veyer D, Wack M, Mandavit M, Garrigou S, Hans S, Bonfils P, Tartour E, Bélec L, Wang-Renault SF, Laurent-Puig P, Mirghani H, Rance B, Taly V, Badoual C, Péré H. HPV circulating tumoral DNA quantification by droplet-based digital PCR: A promising predictive and prognostic biomarker for HPV-associated oropharyngeal cancers. Int J Cancer. 2020;147(4):1222–1227.

91. Akashi K, Sakai T, Fukuoka O, Saito Y, Yoshida M, Ando M, Ito T, Murakami Y, Yamasoba T. Usefulness of circulating tumor DNA by targeting human papilloma virus-derived sequences as a biomarker in p16-positive oropharyngeal cancer. Scientific Reports. 2022;12(1):572.

92. Cocuzza CE, Martinelli M, Sina F, Piana A, Sotgiu G, Dell’Anna T, Musumeci R. Human papillomavirus DNA detection in plasma and cervical samples of women with a recent history of low grade or precancerous cervical dysplasia. PLoS One. 2017;12(11):e0188592.

93. Capone RB, Pai SI, Koch WM, Gillison ML, Danish HN, Westra WH, Daniel R, Shah KV, Sidransky D. Detection and quantitation of human papillomavirus (HPV) DNA in the sera of patients with HPV-associated head and neck squamous cell carcinoma. Clin Cancer Res. 2000;6(11):4171–4175.

94. Ho CM, Yang SS, Chien TY, Huang SH, Jeng CJ, Chang SF. Detection and quantitation of human papillomavirus type 16, 18 and 52 DNA in the peripheral blood of cervical cancer patients. Gynecol Oncol. 2005;99(3):615–621.

95. Hsu KF, Huang SC, Hsiao JR, Cheng YM, Wang SP, Chou CY. Clinical significance of serum human papillomavirus DNA in cervical carcinoma. Obstet Gynecol. 2003;102(6):1344–1351.

96. Mazurek AM, Rutkowski T, Śnietura M, Pigłowski W, Suwiński R, Składowski K. Detection of circulating HPV16 DNA as a biomarker in the blood of patients with human papillomavirus-positive oropharyngeal squamous cell carcinoma. Head Neck. 2019;41(3):632–641.

97. Pornthanakasem W, Shotelersuk K, Termrungruanglert W, Voravud N, Niruthisard S, Mutirangura A. Human papillomavirus DNA in plasma of patients with cervical cancer. BMC Cancer. 2001;1:2.

98. Dahlstrom KR, Li G, Hussey CS, Vo JT, Wei Q, Zhao C, Sturgis EM. Circulating human papillomavirus DNA as a marker for disease extent and recurrence among patients with oropharyngeal cancer. Cancer. 2015;121(19):3455–3464.

99. Reder H, Taferner VF, Wittekindt C, Bräuninger A, Speel EM, Gattenlöhner S, Wolf G, Klussmann JP, Wuerdemann N, Wagner S. Plasma Cell-Free Human Papillomavirus Oncogene E6 and E7 DNA Predicts Outcome in Oropharyngeal Squamous Cell Carcinoma. J Mol Diagn. 2020;22(11):1333–1343.

100. Yang HJ, Liu VW, Tsang PC, Yip AM, Tam KF, Wong LC, Ng TY, Ngan HY. Quantification of human papillomavirus DNA in the plasma of patients with cervical cancer. Int J Gynecol Cancer. 2004;14(5):903–910.

