## Supplementary Materials for "Comparing the Diagnostic Performance of qPCR, ddPCR, and NGS Liquid Biopsies for HPV-Associated Cancers"

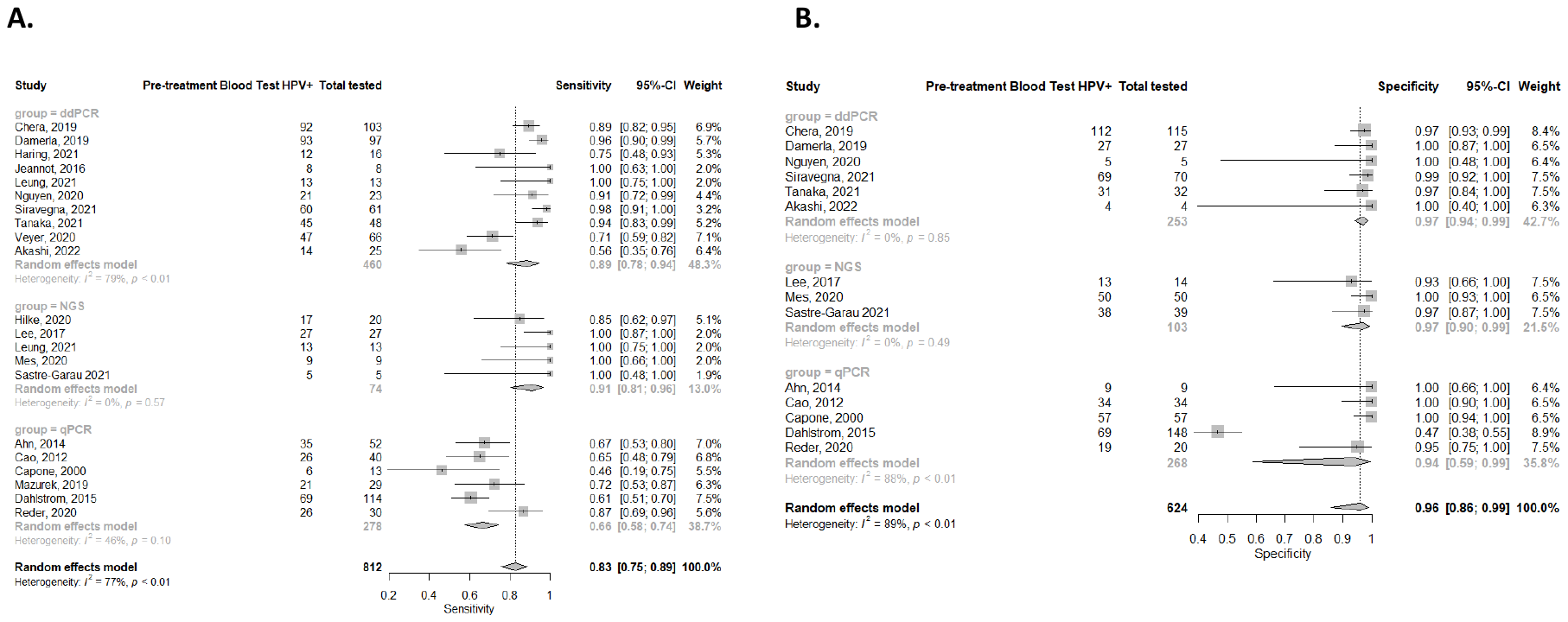

**Figure S1.** Sensitivity (A) and specificity (B) from studies used in the meta-analysis comparing the three assays in HPV+OPSCC.

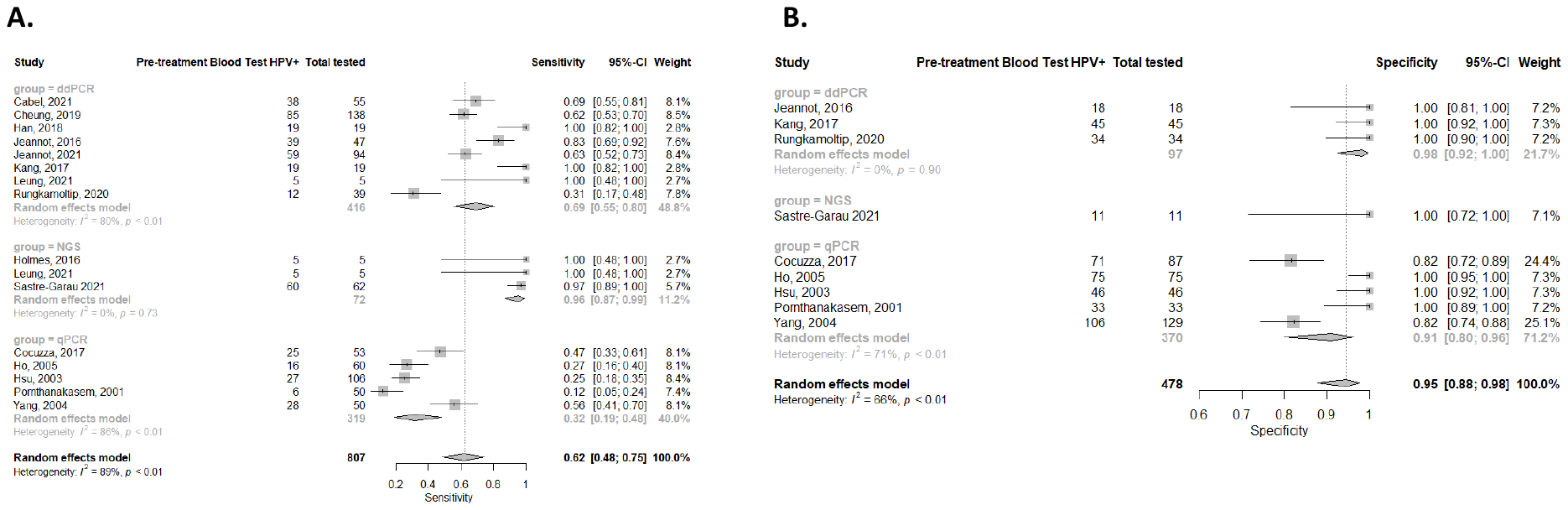

**Figure S2.** Sensitivity (A) and specificity (B) from studies used in the meta-analysis comparing the three assays in HPV+CC.

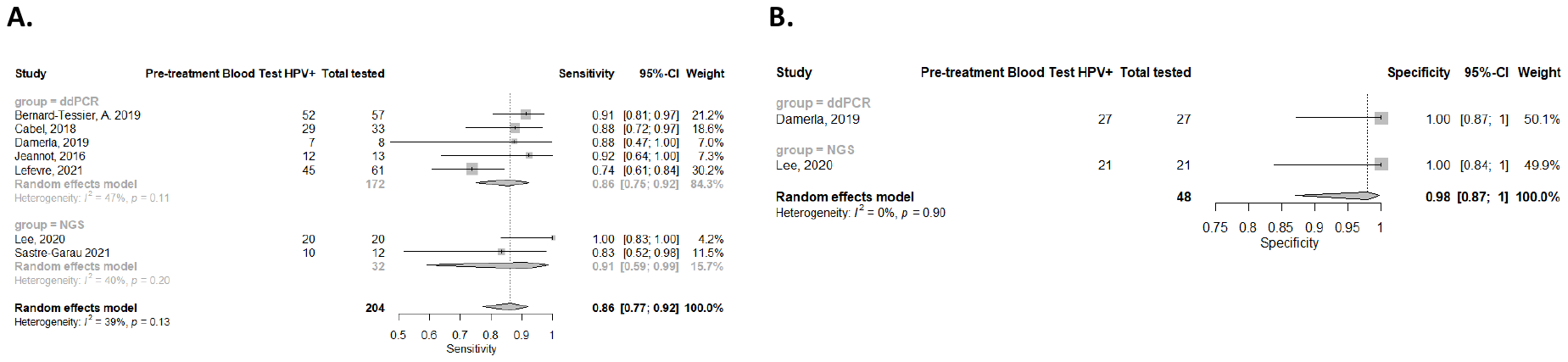

**Figure S3.** Sensitivity (A) and specificity (B) from studies used in the meta-analysis comparing the three assays in HPV+SCCA.

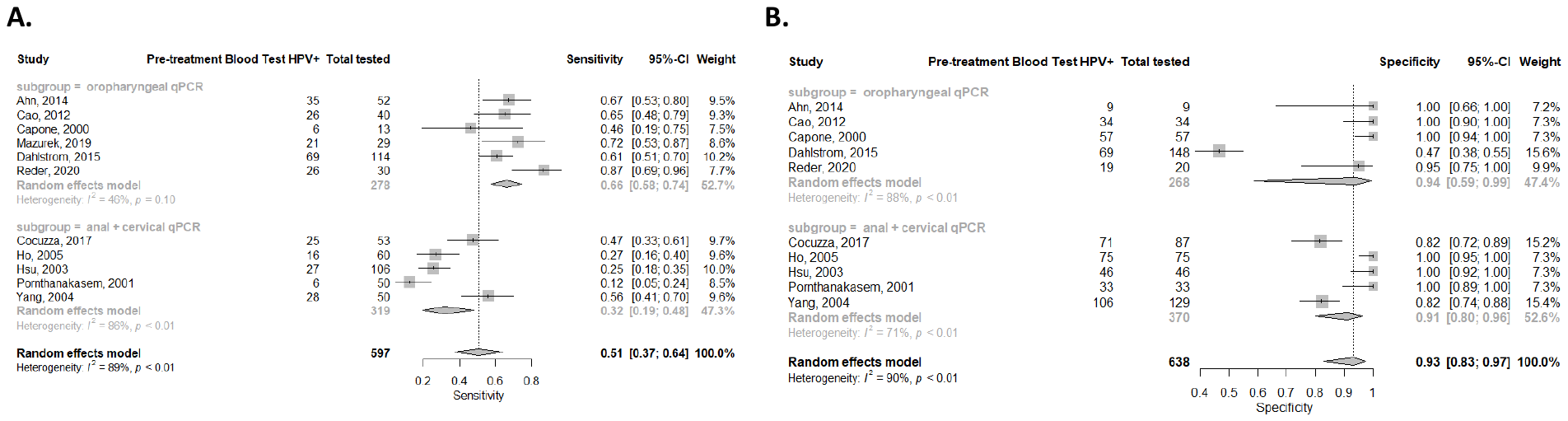

**Figure S4.** Sensitivity (A) and specificity (B) from HPV+OPSCC qPCR studies compared to HPV+CC and HPV+SCCA qPCR studies.

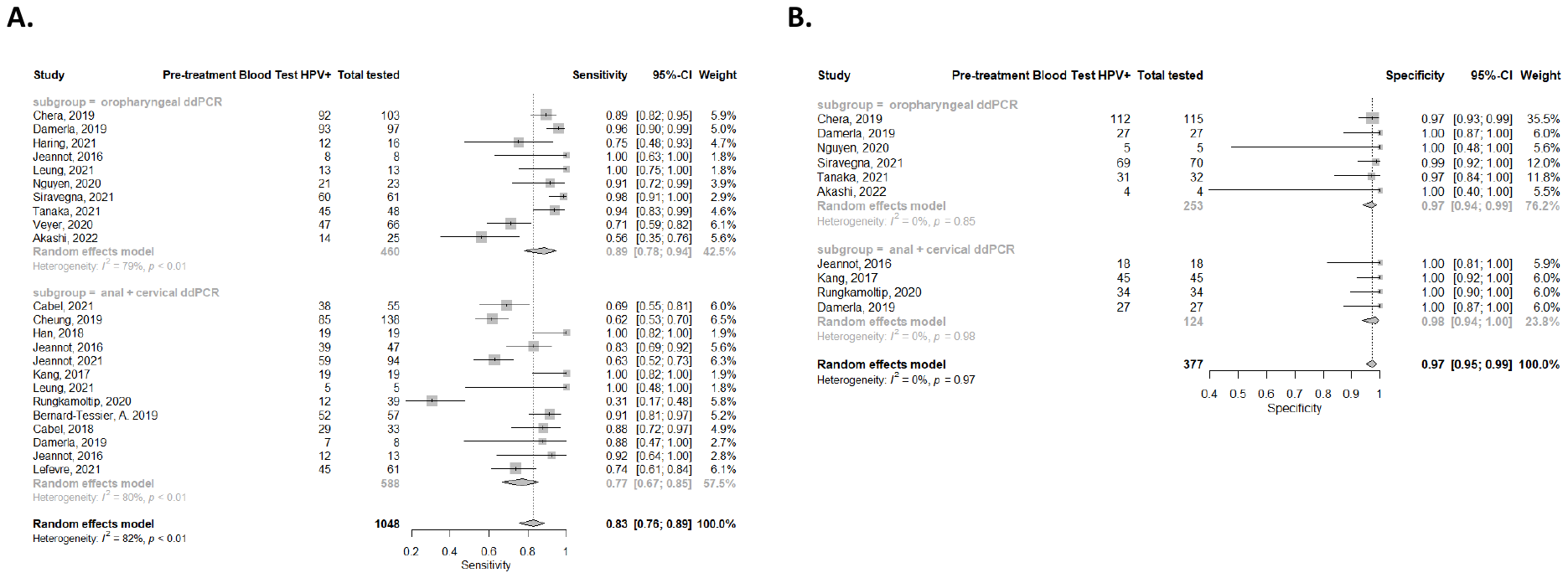

**Figure S5.** Sensitivity (A) and specificity (B) from HPV+OPSCC ddPCR studies compared to HPV+CC and HPV+SCCA ddPCR studies.

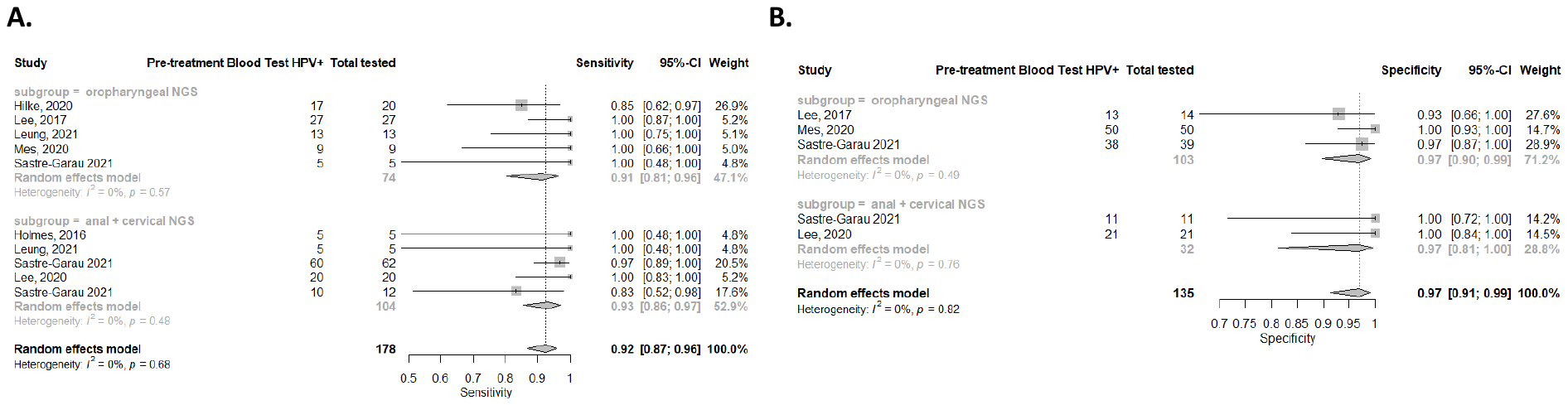

**Figure S6.** Sensitivity (A) and specificity (B) from HPV+OPSCC NGS studies compared to HPV+CC and HPV+SCCA NGS studies.

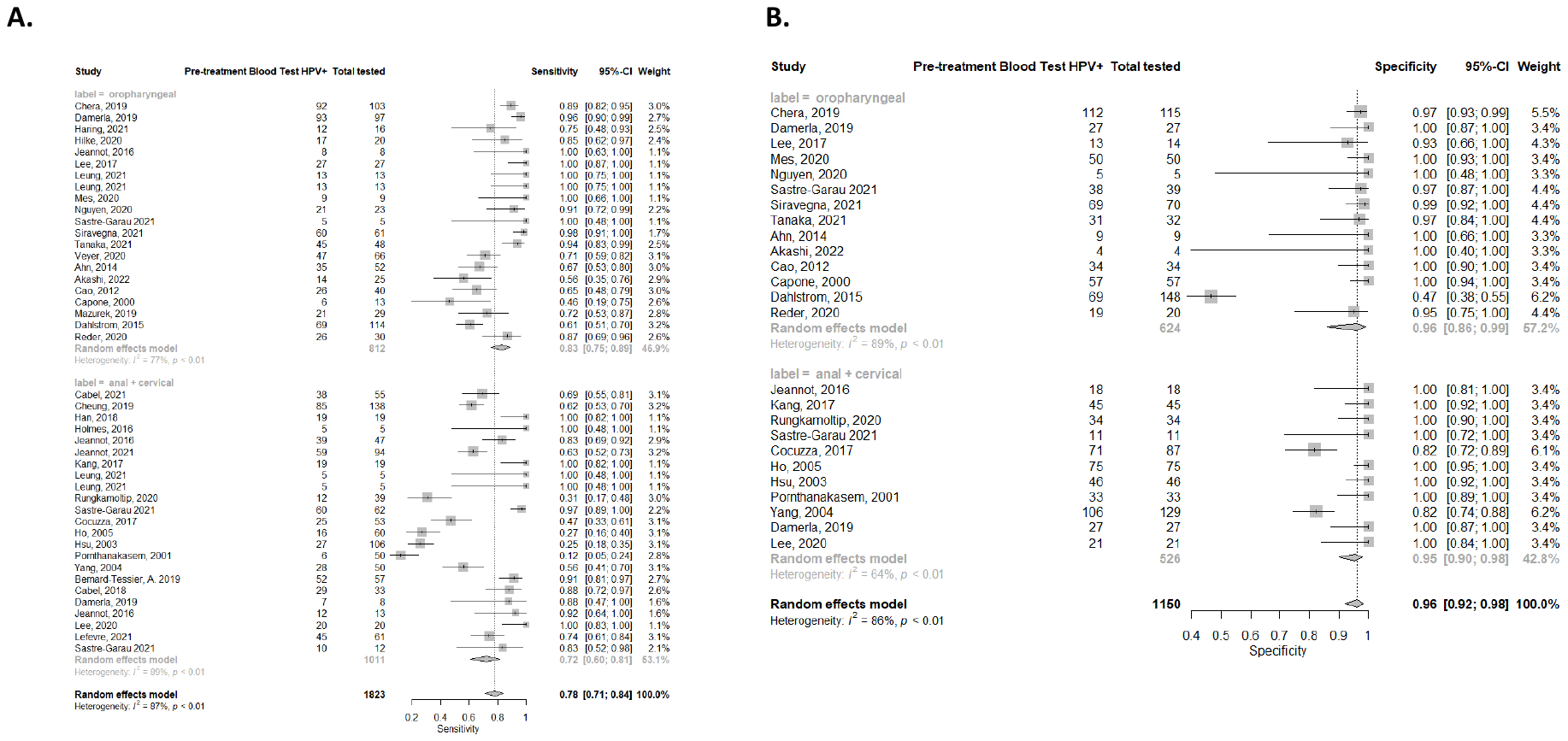

**Figure S7.** Sensitivity (A) and specificity (B) from HPV+OPSCC qPCR, ddPCR, and NGS studies compared to HPV+CC and HPV+SCCA NGS qPCR, ddPCR, and NGS studies.

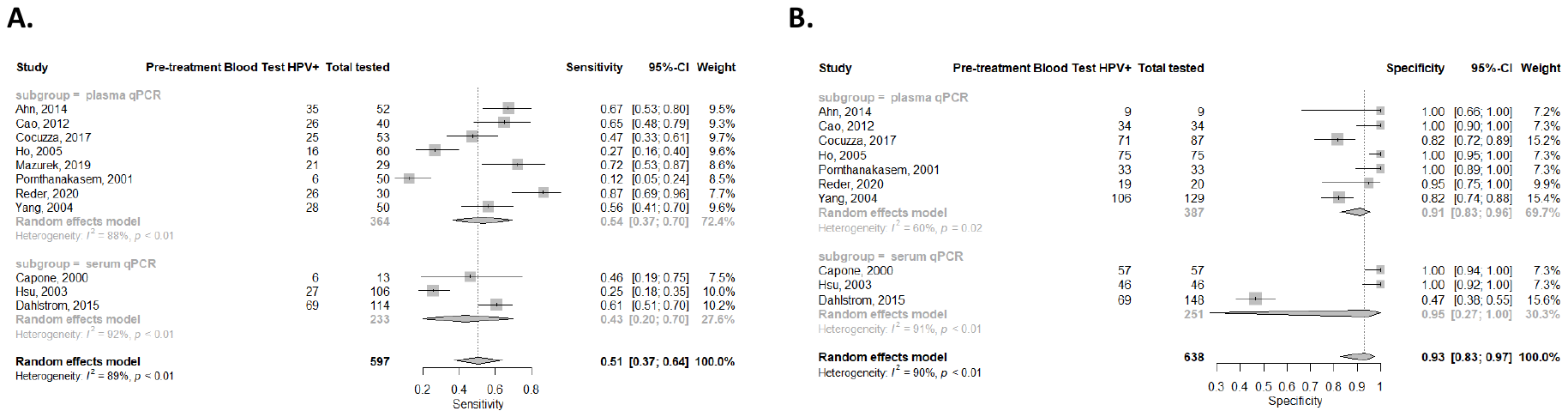

**Figure S8.** Sensitivity (A) and specificity (B) from qPCR studies using serum compared to qPCR studies using plasma. A pooled sensitivity of 0.43 (95% CI: 0.20-0.70) from three serum qPCR studies (*n*=233) was compared to 0.54 (95% CI: 0.37-0.70) from eight plasma qPCR studies (*n*= 364) (p=0.540). A pooled specificity of 0.95 (95% CI: 0.27-1.00) from three qPCR studies (*n*=251) using serum was compared to 0.91 (95% CI: 0.83-0.96) from seven qPCR studies (*n*=387) using plasma (p=0.493).

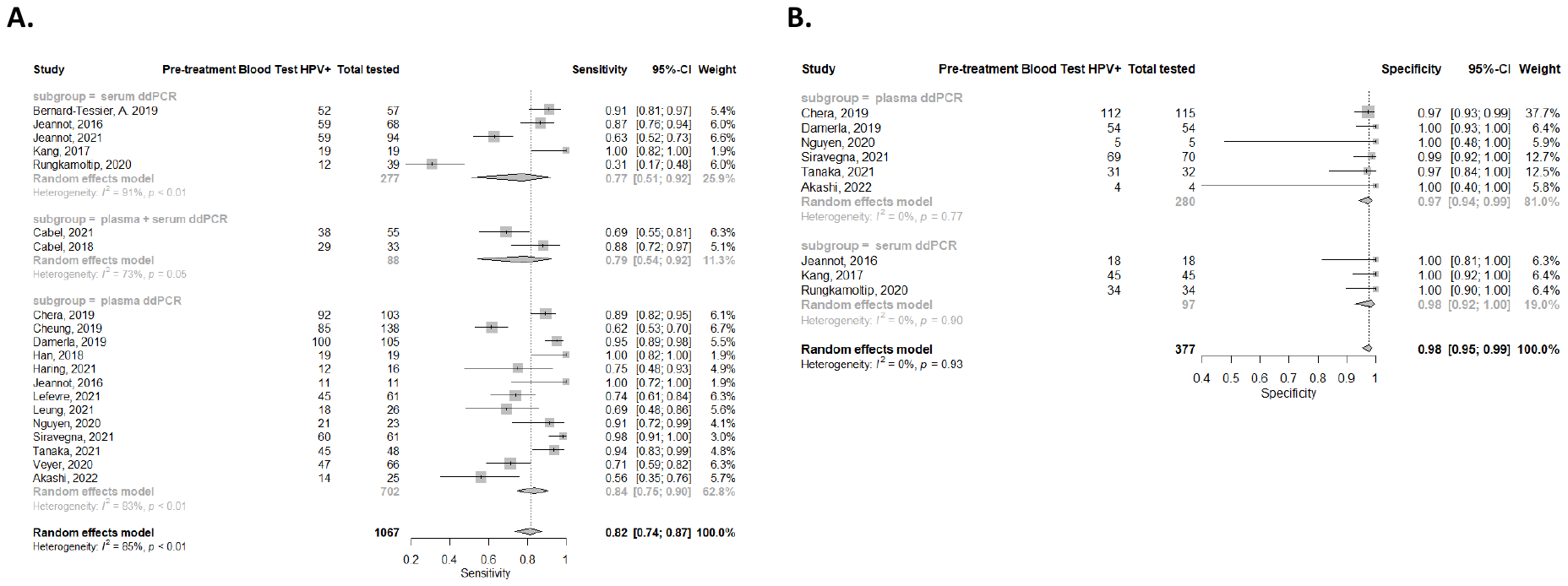

**Figure S9.** Sensitivity (A) and specificity (B) from ddPCR studies using serum compared to ddPCR studies using plasma and serum or plasma alone. A pooled sensitivity of 0.77 (95% CI: 0.51-0.92) from five serum ddPCR studies (*n*=277) was compared to 0.79 (95% CI: 0.54-0.92) from two ddPCR studies (*n*=88) that used both plasma and serum samples (p=0.845). A pooled sensitivity of 0.84 (95% CI: 0.75-0.90) from 13 ddPCR studies (*n*=702) using plasma was then compared to the ddPCR studies using both plasma and serum (p=0.674). A pooled specificity of 0.98 (95% CI: 0.92-1.00) from three ddPCR studies using serum (*n*=97) was compared to 0.97 (95% CI: 0.94-0.99) from six ddPCR studies (*n*=280) using plasma (p=0.555).

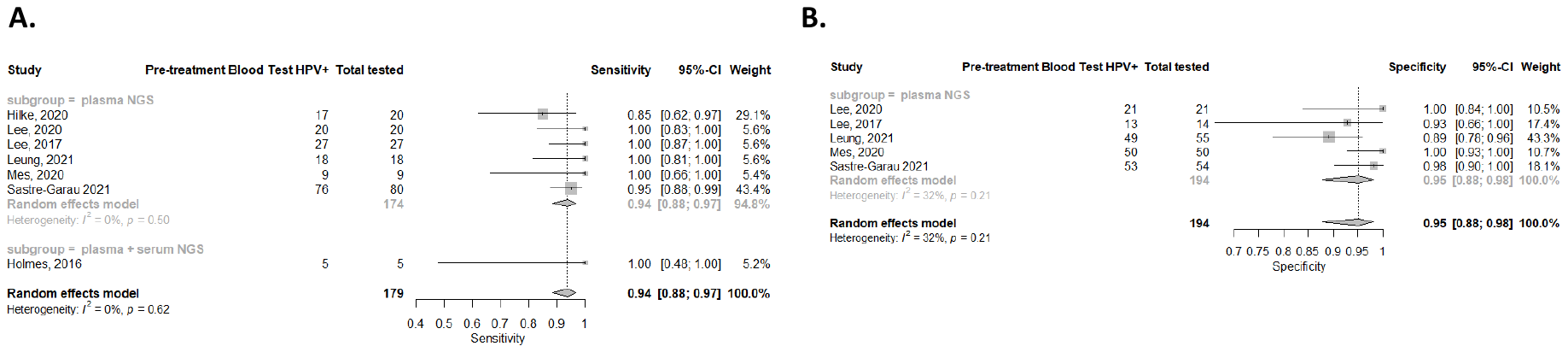

**Figure S10.** Sensitivity (A) and specificity (B) from NGS studies using serum compared to NGS studies using plasma and serum. A pooled sensitivity of 0.94 (95% CI: 0.88-0.97) from six NGS studies (*n*=174) using plasma was compared to 1.00 (95% CI: 0.48-1.00) from one NGS study (*n*=5) using both plasma and serum (p=0.832). Specificity regression was unavailable for the NGS studies because of small sample size.

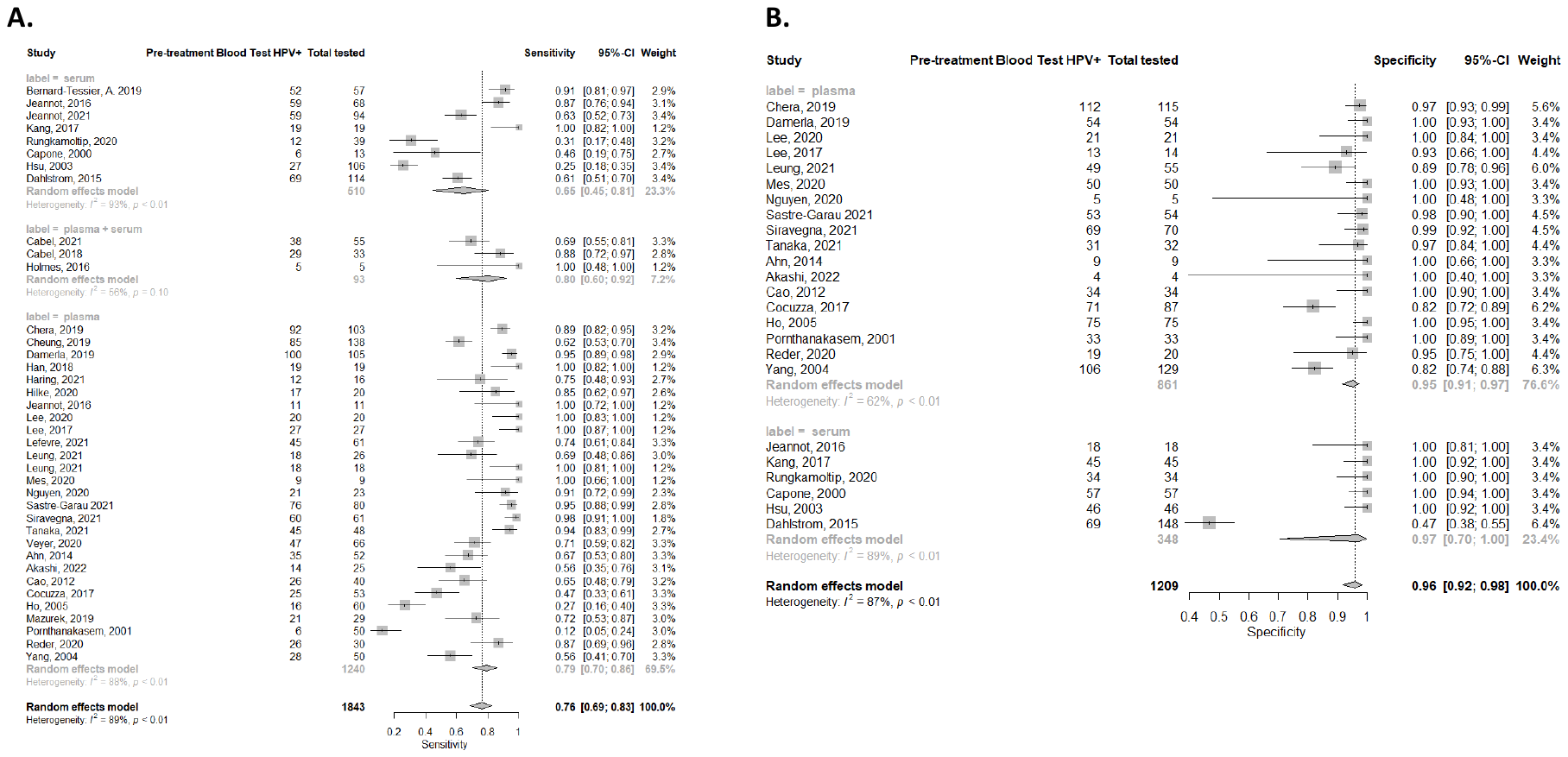

**Figure S11.** Sensitivity (A) and specificity (B) from qPCR, ddPCR, and NGS studies overall using serum compared to qPCR, ddPCR, and NGS studies overall using plasma and serum or plasma alone.

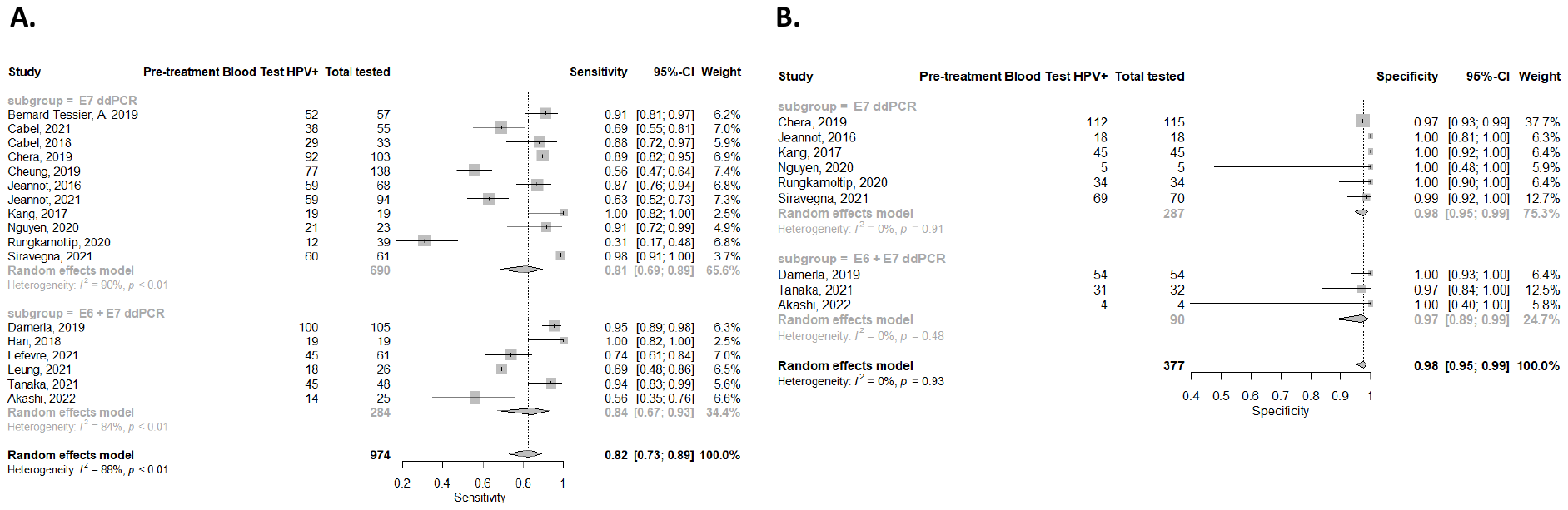

**Figure S12.** Sensitivity (A) and specificity (B) from ddPCR studies using probes for E7 target gene compared to ddPCR studies using probes for both E6 and E7 genes.

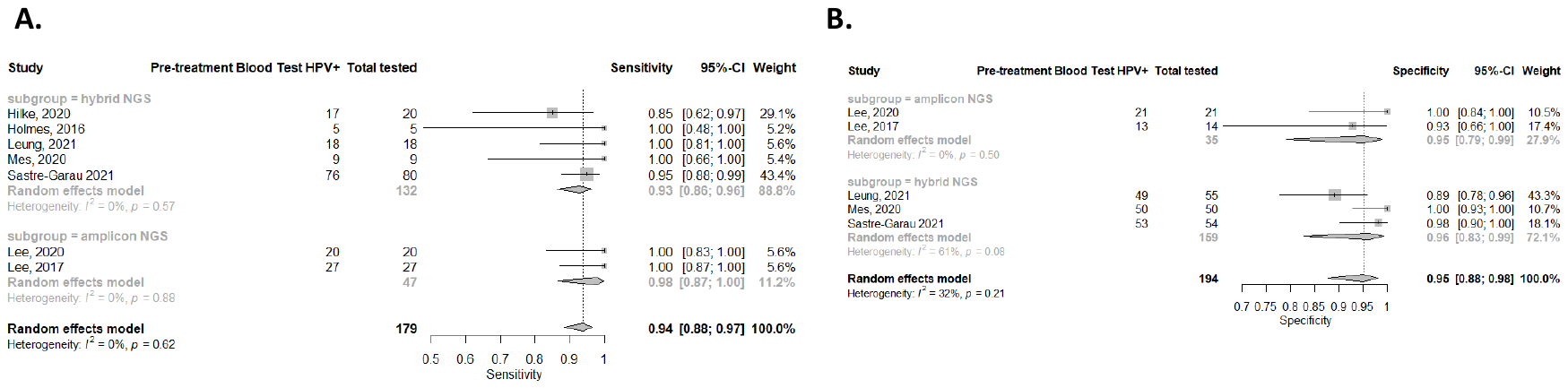

**Figure S13.** Sensitivity (A) and specificity (B) from NGS studies using an amplicon-based approach compared to NGS studies using a hybrid capture-based approach.

**Supplementary Methods**

**Complete Embase Search**

**head and neck cancer**

'head and neck tumor'/exp OR 'nasopharynx tumor'/exp OR 'hypopharynx tumor'/exp OR 'oropharynx tumor'/exp OR oropharynx:ti,ab OR 'mouth tumor'/exp OR 'nose tumor'/exp OR 'paranasal sinus tumor'/exp OR 'maxilla sinus cancer'/exp OR 'ear tumor'/exp OR 'head and neck cancer':ti,ab OR 'head and neck neoplasm*':ti,ab OR 'head and neck tumour':ti,ab OR 'head and neck tumor':ti,ab OR HNSCC:ti,ab OR 'cervicofacial cancer':ti,ab OR 'ear nose throat cancer':ti,ab OR 'ENT cancer':ti,ab OR 'head neck cancer':ti,ab OR 'ORL cancer':ti,ab OR 'otorhinolaryngeal cancer':ti,ab OR 'otorhinolaryngologic cancer':ti,ab OR 'otorhinolaryngological cancer':ti,ab OR 'facial cancer':ti,ab OR ((cancer OR carcinoma OR tumor OR tumour OR neoplasm) NEAR/3 (larynx OR laryngeal OR nasopharynx OR nasopharyngeal OR hypopharynx OR hypopharyngeal OR oropharynx OR oropharyngeal OR tracheal OR trachea)):ti,ab OR ((cancer OR carcinoma OR tumor OR tumour OR neoplasm) NEAR/3 ('tonsil*' OR 'tongue' OR 'base of tongue' OR 'mouth' OR 'oral cavit*')):ti,ab OR ((cancer OR carcinoma OR tumor OR tumour OR neoplasm) NEAR/3 (nose OR nasal)):ti,ab OR ((cancer OR carcinoma OR tumor OR tumour OR neoplasm) NEAR/3 ('maxilla sinus' OR 'paranasal sinus' OR 'ethmoid sinus' OR 'sphenoid sinus' OR 'frontal sinus')):ti,ab OR ((cancer OR carcinoma OR tumor OR tumour OR neoplasm) NEAR/3 (ear OR 'temporal bone')):ti,ab OR ((cancer OR carcinoma OR tumor OR tumour OR neoplasm) NEAR/3 (face OR facial)):ti,ab

**393,001**

**Anal carcinoma**

'anus cancer'/exp OR 'anus carcinoma'/exp OR 'anal canal carcinoma':ti,ab OR 'anal carcinoma':ti,ab OR 'anal intraepithelial neoplasia':ti,ab OR 'anus carcinoma':ti,ab OR 'perianal carcinoma':ti,ab OR 'anal squamous cell carcinoma':ti,ab OR 'anal cancer':ti,ab

10,152

**Cervical carcinoma**

'uterine cervix cancer'/exp OR 'uterine cervix cancer':ti,ab OR 'cervical cancer':ti,ab OR 'cervix ca':ti,ab OR 'cervix cancer':ti,ab OR 'recurrent cervix cancer':ti,ab OR 'uterine cervix cancer':ti,ab OR 'uterine cervix':ti,ab OR 'recurrent cervix malignancy':ti,ab OR 'cervix uteri cancer':ti,ab OR 'cervix uterus cancer':ti,ab OR 'neoplasma cervicis recurrens':ti,ab OR 'neoplasma cervicis uteri recurrens':ti,ab OR 'uterine cervical cancer':ti,ab OR 'recurrent uterine cervix cancer':ti,ab OR 'recurrent uterine cervix malignancy':ti,ab OR 'uterus cervix cancer':ti,ab OR 'cervical carcinoma*':ti,ab

140,755

1 OR 2 OR 3 = 533,680

**HPV**

'Papillomaviridae'/exp OR 'papillomavirus infection'/exp OR HPV:ti,ab OR 'HPV+':ti,ab OR 'p16+':ti,ab OR 'p16':ti,ab OR 'HPV-16':ti,ab OR 'HPV-18':ti,ab OR 'papilloma virus*':ti,ab OR papillomavirus*:ti,ab OR 'Circulating HPV DNA':ti,ab OR 'Human papillomavirus (HPV)–associated carcinomas':ti,ab

119,195

4 AND 5 = 50,900

'liquid biopsy'/exp OR 'high throughput sequencing'/exp OR 'ngs analysis (next generation sequence analysis)':ti,ab OR 'ngs-based':ti,ab OR 'high through-put nucleotide sequencing':ti,ab OR 'high through-put sequence analysis':ti,ab OR 'high through-put sequencing':ti,ab OR 'high throughput nucleotide sequence analysis':ti,ab OR 'high throughput nucleotide sequencing':ti,ab OR 'high throughput sequence analysis':ti,ab OR 'high throughput sequencing':ti,ab OR 'high-throughput nucleotide sequencing':ti,ab OR 'next generation sequence analysis':ti,ab OR 'next generation sequencing':ti,ab OR 'next generation sequencing technology':ti,ab OR 'next-gen sequence analysis':ti,ab OR 'next-gen sequencing':ti,ab OR 'liquid biops*':ti,ab OR 'NGS-based':ti,ab

145,421

'polymerase chain reaction system'/exp OR 'droplet digital polymerase chain reaction'/exp OR 'dd-pcr':ti,ab OR 'ddpcr':ti,ab OR 'digital droplet pcr':ti,ab OR 'digital droplet polymerase chain reaction':ti,ab OR 'droplet digital pcr':ti,ab OR 'droplet digital polymerase chain reaction':ti,ab OR 'digital polymerase chain reaction':ti,ab OR 'digital PCR assay*':ti,ab OR 'polymerase chain reaction system*':ti,ab

'real time polymerase chain reaction'/exp OR 'real time polymerase chain reaction':ti,ab OR qRT-PCR:ti,ab OR qRTPCR:ti,ab OR 'quantitative PCR':ti,ab OR 'quantitative polymerase chain reaction':ti,ab OR 'real time PCR':ti,ab OR 'real-time PCR':ti,ab OR 'real time qPCR':ti,ab OR 'real time quantitative PCR':ti,ab OR 'real time quantitative polymerase chain reaction':ti,ab OR 'real-time polymerase chain reaction':ti,ab OR 'realtime PCR':ti,ab OR 'realtime polymerase chain reaction':ti,ab OR 'realtime quantitative PCR':ti,ab OR 'realtime quantitative polymerase chain reaction':ti,ab OR 'RT PCR':ti,ab OR 'RT-qPCR':ti,ab OR 'RTQ-PCR':ti,ab OR 'RTqPCR':ti,ab OR 'Kinetic Polymerase Chain Reaction':ti,ab OR 'Kinetic PCR':ti,ab

28,481

7 OR 8 = 170,837

6 AND 9 = 916

'circulating tumor DNA'/exp OR 'circulating tumor DNA':ti,ab OR 'cell-free DNA':ti,ab OR 'circulating viral DNA':ti,ab OR 'Circulating HPV DNA':ti,ab OR 'Plasma HPV DNA':ti,ab OR 'plasma human papillomavirus (HPV) DNA':ti,ab OR 'circulating tumor cell*':ti,ab OR 'plasma circulating tumor HPV':ti,ab

25,301

10 AND 11 = 99

(34108183 OR 28809864 OR 32687856):ui OR L631620514 OR L2002633207 OR (31088830 OR 31756275 OR 32269293 OR 28899967 OR 34210686 OR 30010779 OR 30913520 OR 32017652 OR 30054279 OR 30504426):ui OR L627280121 OR L2005928548 OR L616970469 OR L2013349898 OR L627768624

**Complete MEDLINE Search**

**Head and neck cancer**

exp Head and Neck Neoplasms/ OR exp Nasopharyngeal Neoplasms/ OR exp Hypopharyngeal Neoplasms/ OR exp Oropharyngeal Neoplasms/ OR exp Tonsillar Neoplasms/ OR exp Tongue Neoplasms/ OR exp Mouth Neoplasms/ OR exp Laryngeal Neoplasms/ OR exp Nose Neoplasms/ OR exp Paranasal Sinus Neoplasms/ OR exp Maxillary Sinus Neoplasms/ OR exp Ear Neoplasms/ OR "head and neck cancer".ti,ab. OR "head and neck neoplasm*".ti,ab. OR "head and neck tumour".ti,ab. OR "head and neck tumor".ti,ab. OR HNSCC.ti,ab. OR "cervicofacial cancer".ti,ab. OR "ear nose throat cancer".ti,ab. OR "ENT cancer".ti,ab. OR "head neck cancer".ti,ab. OR "ORL cancer".ti,ab. OR "otorhinolaryngeal cancer".ti,ab. OR "otorhinolaryngologic cancer".ti,ab. OR "otorhinolaryngological cancer".ti,ab. OR "facial cancer".ti,ab. OR ((cancer OR carcinoma OR tumor OR tumour OR neoplasm) ADJ3 (larynx OR laryngeal OR nasopharynx OR nasopharyngeal OR hypopharynx OR hypopharyngeal OR oropharynx OR oropharyngeal OR tracheal OR tracheal)).ti,ab. OR ((cancer OR carcinoma OR tumor OR tumour OR neoplasm) ADJ3 ("tonsil*" OR "tongue" OR "base of tongue" OR "mouth" OR "oral cavit*")).ti,ab. OR ((cancer OR carcinoma OR tumor OR tumour OR neoplasm) ADJ3 (nose OR nasal)).ti,ab. OR ((cancer OR carcinoma OR tumor OR tumour OR neoplasm) ADJ3 ("maxilla sinus" OR "paranasal sinus" OR "ethmoid sinus" OR "sphenoid sinus" OR "frontal sinus")).ti,ab. OR ((cancer OR carcinoma OR tumor OR tumour OR neoplasm) ADJ3 (ear OR "temporal bone")).ti,ab. OR ((cancer OR carcinoma OR tumor OR tumour OR neoplasm) ADJ3 (face OR facial)).ti,ab.

190,914

**Anal carcinoma**

Exp Anus neoplasms/ OR "anal canal carcinoma".ti,ab. OR "anal carcinoma".ti,ab. OR "anal intraepithelial neoplasia".ti,ab. OR "anus carcinoma".ti,ab. OR "perianal carcinoma".ti,ab. OR "anal squamous cell carcinoma".ti,ab. OR "anal cancer".ti,ab.

7727

**Cervical carcinoma**

Exp Uterine Cervical Neoplasms/ OR "uterine cervix cancer".ti,ab. OR "cervical cancer".ti,ab. OR "cervix ca".ti,ab. OR "cervix cancer".ti,ab. OR "recurrent cervix cancer".ti,ab. OR "uterine cervix cancer".ti,ab. OR "uterine cervix".ti,ab. OR "recurrent cervix malignancy".ti,ab. OR "cervix uteri cancer".ti,ab. OR "cervix uterus cancer".ti,ab. OR "neoplasma cervicis recurrens".ti,ab. OR "neoplasma cervicis uteri recurrens".ti,ab. OR "uterine cervical cancer".ti,ab. OR "recurrent uterine cervix cancer".ti,ab. OR "recurrent uterine cervix malignancy".ti,ab. OR "uterus cervix cancer".ti,ab. OR "cervical carcinoma*".ti,ab.

101,525

1 OR 2 OR 3 =297,557

**HPV**

Exp Papillomaviridae/ OR Exp Papillomavirus Infections/ OR HPV.ti,ab. OR "HPV+".ti,ab. OR "p16+".ti,ab. OR "p16".ti,ab. OR "HPV-16".ti,ab. OR "HPV-18".ti,ab. OR "papilloma virus*".ti,ab. OR papillomavirus*.ti,ab. OR "Circulating HPV DNA".ti,ab. OR "Human papillomavirus (HPV)–associated carcinomas".ti,ab.

81,545

4 & 5 = 35,481

**Testing**

**NGS**

Exp Liquid Biopsy/ OR Exp High-Throughput Nucleotide Sequencing/ OR "ngs analysis (next generation sequence analysis)".ti,ab. OR "ngs-based".ti,ab. OR "high through-put nucleotide sequencing".ti,ab. OR "high through-put sequence analysis".ti,ab. OR "high through-put sequencing".ti,ab. OR "high throughput nucleotide sequence analysis".ti,ab. OR "high throughput nucleotide sequencing".ti,ab. OR "high throughput sequence analysis".ti,ab. OR "high throughput sequencing".ti,ab. OR "high-throughput nucleotide sequencing".ti,ab. OR "next generation sequence analysis".ti,ab. OR "next generation sequencing".ti,ab. OR "next generation sequencing technology".ti,ab. OR "next-gen sequence analysis".ti,ab. OR "next-gen sequencing".ti,ab. OR "liquid biops*".ti,ab.

82,698

**ddPCR**

"dd-pcr".ti,ab. OR "ddpcr".ti,ab. OR "digital droplet pcr".ti,ab. OR "digital droplet polymerase chain reaction".ti,ab. OR "droplet digital pcr".ti,ab. OR "droplet digital polymerase chain reaction".ti,ab. OR "digital polymerase chain reaction".ti,ab. OR "digital PCR assay*".ti,ab. OR "polymerase chain reaction system*".ti,ab.

exp Real-Time Polymerase Chain Reaction/ OR "real time polymerase chain reaction".ti,ab. OR qRT-PCR.ti,ab. OR qRTPCR.ti,ab. OR "quantitative PCR".ti,ab. OR "quantitative polymerase chain reaction".ti,ab. OR "real time PCR".ti,ab. OR "real-time PCR".ti,ab. OR "real time qPCR".ti,ab. OR "real time quantitative PCR".ti,ab. OR "real time quantitative polymerase chain reaction".ti,ab. OR "real-time polymerase chain reaction":ti,,ab OR "realtime PCR".ti,ab. OR "realtime polymerase chain reaction".ti,ab. OR "realtime quantitative PCR".ti,ab. OR "realtime quantitative polymerase chain reaction".ti,ab. OR "RT PCR".ti,ab OR "RT-qPCR".ti,ab. OR "RTQ-PCR".ti,ab. OR "RTqPCR".ti,ab. OR "Kinetic Polymerase Chain Reaction".ti,ab. OR "Kinetic PCR".ti,ab.

28,401

7 OR 8 = 85,842

6 AND 9 = 337

Exp circulating tumor DNA/ OR "circulating tumor DNA".ti,ab. OR "cell-free DNA".ti,ab. OR "circulating viral DNA".ti,ab. OR "Circulating HPV DNA".ti,ab. OR "Plasma HPV DNA".ti,ab. OR "plasma human papillomavirus (HPV) DNA".ti,ab. OR "Circulating DNA".ti,ab. OR "circulating tumor cell*".ti,ab. OR "plasma circulating tumor HPV".ti,ab.

13,818

10 AND 11 = 39

(34108183 OR 28809864 OR 32363162 OR 29263809 OR 32687856)

31756275 OR 32269293 OR 28899967 OR 33540527 OR 27917295 OR 34210686 OR 34194620 OR 30010779 OR 31485558 OR 30913520 OR 32017652 OR 31088830 OR 30054279 OR 30504426

**Web of Science – All Databases**

**Head and neck cancer**

"head and neck cancer" OR "head and neck neoplasm*" OR "head and neck tumour" OR "head and neck tumor" OR HNSCC OR "cervicofacial cancer" OR "ear nose throat cancer" OR "ENT cancer" OR "head neck cancer" OR "ORL cancer" OR "otorhinolaryngeal cancer" OR "otorhinolaryngologic cancer" OR "otorhinolaryngological cancer" OR "facial cancer" OR ((cancer OR carcinoma OR tumor OR tumour OR neoplasm) NEAR/3 (larynx OR laryngeal OR nasopharynx OR nasopharyngeal OR hypopharynx OR hypopharyngeal OR oropharynx OR oropharyngeal OR tracheal OR tracheal)) OR ((cancer OR carcinoma OR tumor OR tumour OR neoplasm) NEAR/3 ("tonsil*" OR "tongue" OR "base of tongue" OR "mouth" OR "oral cavit*")) OR ((cancer OR carcinoma OR tumor OR tumour OR neoplasm) NEAR/3 (nose OR nasal)) OR ((cancer OR carcinoma OR tumor OR tumour OR neoplasm) NEAR/3 ("maxilla sinus" OR "paranasal sinus" OR "ethmoid sinus" OR "sphenoid sinus" OR "frontal sinus")) OR ((cancer OR carcinoma OR tumor OR tumour OR neoplasm) NEAR/3 (ear OR "temporal bone")) OR ((cancer OR carcinoma OR tumor OR tumour OR neoplasm) NEAR/3 (face OR facial))

296,511

**Anal carcinoma**

"Anus neoplasms" OR "anal canal carcinoma" OR "anal carcinoma" OR "anal intraepithelial neoplasia" OR "anus carcinoma" OR "perianal carcinoma" OR "anal squamous cell carcinoma" OR "anal cancer"

12,280

**Cervical carcinoma**

"Uterine Cervical Neoplasms" OR "uterine cervix cancer" OR "cervical cancer" OR "cervix ca" OR "cervix cancer" OR "recurrent cervix cancer" OR "uterine cervix" OR "recurrent cervix malignancy" OR "cervix uteri cancer" OR "cervix uterus cancer" OR "neoplasma cervicis recurrens" OR "neoplasma cervicis uteri recurrens" OR "uterine cervical cancer" OR " recurrent uterine cervix cancer" OR "recurrent uterine cervix malignancy" OR "uterus cervix cancer" OR "cervical carcinoma*"

208,044

1 OR 2 OR 3 = 500,491

**HPV**

Papillomaviridae OR "Papillomavirus Infections" OR HPV OR HPV+ OR p16+ OR p16 OR HPV-16 OR HPV-18 OR "papilloma virus*" OR papillomavirus* OR "Circulating HPV DNA" OR "Human papillomavirus (HPV)–associated carcinomas"

147,280

4 AND 5 = 59,224

**Testing**

"ngs analysis (next generation sequence analysis)" OR "ngs-based" OR "high through-put nucleotide sequencing" OR "high through-put sequence analysis" OR "high through-put sequencing" OR "high throughput nucleotide sequence analysis" OR "high throughput nucleotide sequencing" OR "high throughput sequence analysis" OR "high throughput sequencing" OR "high-throughput nucleotide sequencing" OR "next generation sequence analysis" OR "next generation sequencing" OR "next generation sequencing technology" OR "next-gen sequence analysis" OR "next-gen sequencing" OR "liquid biops*"

1,110,580

"droplet digital polymerase chain reaction" OR "dd-pcr" OR "ddpcr" OR "digital droplet pcr" OR "digital droplet polymerase chain reaction" OR "droplet digital pcr" OR "droplet digital polymerase chain reaction" OR "digital polymerase chain reaction"

"real time polymerase chain reaction" OR qRT-PCR OR qRTPCR OR "quantitative PCR" OR "quantitative polymerase chain reaction" OR "real time PCR" OR "real-time PCR" OR "real time qPCR" OR "real time quantitative PCR" OR "real time quantitative polymerase chain reaction" OR "real-time polymerase chain reaction":ti,,ab OR "realtime PCR" OR "realtime polymerase chain reaction" OR "realtime quantitative PCR" OR "realtime quantitative polymerase chain reaction" OR "RT PCR".ti,ab OR "RT-qPCR" OR "RTQ-PCR" OR "RTqPCR" OR "Kinetic Polymerase Chain Reaction" OR "Kinetic PCR"

7,266

7 OR 8 = 1,116,413

6 AND 9 = 568

"circulating tumor DNA" OR "cell-free DNA" OR "circulating viral DNA" OR "Circulating HPV DNA" OR "Plasma HPV DNA" OR "plasma human papillomavirus (HPV) DNA" OR "Circulating DNA" OR "circulating tumor cell*" OR "plasma circulating tumor HPV"

31,837

10 AND 11 = 77
